# Large-scale multi-omic analysis identifies noncoding somatic driver mutations and nominates *ZFP36L2* as a driver gene for pancreatic ductal adenocarcinoma

**DOI:** 10.1101/2024.09.22.24314165

**Authors:** Jun Zhong, Aidan O’Brien, Minal Patel, Daina Eiser, Michael Mobaraki, Irene Collins, Li Wang, Konnie Guo, ThucNhi TruongVo, Ashley Jermusyk, Maura O’Neill, Courtney D. Dill, Andrew D. Wells, Michelle E. Leonard, James A. Pippin, Struan F.A. Grant, Tongwu Zhang, Thorkell Andresson, Katelyn E. Connelly, Jianxin Shi, H. Efsun Arda, Jason W. Hoskins, Laufey T. Amundadottir

## Abstract

Identification of somatic driver mutations in the noncoding genome remains challenging. To comprehensively characterize noncoding driver mutations for pancreatic ductal adenocarcinoma (PDAC), we first created genome-scale maps of accessible chromatin regions (ACRs) and histone modification marks (HMMs) in pancreatic cell lines and purified pancreatic acinar and duct cells. Integration with whole-genome mutation calls from 506 PDACs revealed 314 ACRs/HMMs significantly enriched with 3,614 noncoding somatic mutations (NCSMs). Functional assessment using massively parallel reporter assays (MPRA) identified 178 NCSMs impacting reporter activity (19.45% of those tested). Focused luciferase validation confirmed negative effects on gene regulatory activity for NCSMs near *CDKN2A* and *ZFP36L2*. For the latter, CRISPR interference (CRISPRi) further identified *ZFP36L2* as a target gene (16.0 – 24.0% reduced expression, *P* = 0.023-0.0047) with disrupted KLF9 binding likely mediating the effect. Our integrative approach provides a catalog of potentially functional noncoding driver mutations and nominates *ZFP36L2* as a PDAC driver gene.

## Introduction

Driven mainly by diagnosis at a late stage and a lack of effective treatment modalities, pancreatic cancer is characterized by an overall 5-year survival rate of only 13% and is predicted to become the second leading cause of cancer-related deaths in the U.S. by the end of this decade^1–3^. This highlights an urgent need for further research into the etiology of PDAC to improve outcomes for this cancer of unmet need. Cancer develops due to somatic and germline genetic factors exerting their influence within the cellular context of environmental factors^4^. Identification of driver mutations can significantly enhance our understanding of tumorigenesis and facilitate precision oncology^5^. Thanks to large-scale whole exome sequencing studies, protein-coding somatic driver mutations for PDAC (e.g. in *KRAS*, *TP53*, *SMAD4*, *CDKN2A*) have been well characterized^6–8^. The discovery of recurrent noncoding mutations in *cis*-regulatory elements (CREs) implicated in the expression regulation of genes associated with oncogenesis, such as in the *TERT* promoter^9,10^, *TAL1* super-enhancer^11^, and *PAX5* enhancers^12^, has highlighted the emerging role of somatic mutations in noncoding regulatory regions as key contributors to carcinogenesis. Although most somatic mutations occur in noncoding regions of the genome, identifying driver mutations and their target genes has remained largely elusive.

NCSMs can disrupt transcription factor binding sites that impact the activity of gene regulatory elements including promoters and enhancers^13^. These elements are often highly tissue or cell type-specific, with publicly available annotations of PDAC-relevant elements limited in scope and sample size^14,15^. With this in mind, we generated genome-wide maps of ACRs and HMMs in pancreatic tumor- and normal-derived pancreatic cell lines, as well as from pancreatic acinar and ductal cell populations purified using flow cytometry. We integrated these elements with NCSMs identified in 506 PDAC genomes^16–18^ to identify potential drivers, while adjusting for background mutational rates. To characterize the functional consequences of NCSMs, we applied MPRA^19^ to identify mutations associated with altered transcriptional regulatory activity. Subsequently, we used genome-scale high-resolution promoter-focused Capture-C (PCC)^20^ and H3K27ac HiChIP-seq (HiChIP)^21^ to identify chromatin interactions that link NCSMs to putative target genes. Our work provides new insights into the potential role of NCSMs in PDAC development and establishes an effective framework for applying a multi-omics approach to unravel complex CREs in their relation to cancer biology.

## Results

We leveraged an integrative multi-omics approach, using genome, epigenome, chromatin interactome, and transcriptome datasets for PDAC, to identify candidate noncoding driver mutations for further analysis. Putative driver mutations were then characterized by MPRA, investigation of disrupted transcription factor binding sites, and alternative splicing analysis that ultimately prioritized a set of NCSMs most likely to represent noncoding driver mutations (study design outlined in Fig. 1a).

**Fig. 1.**
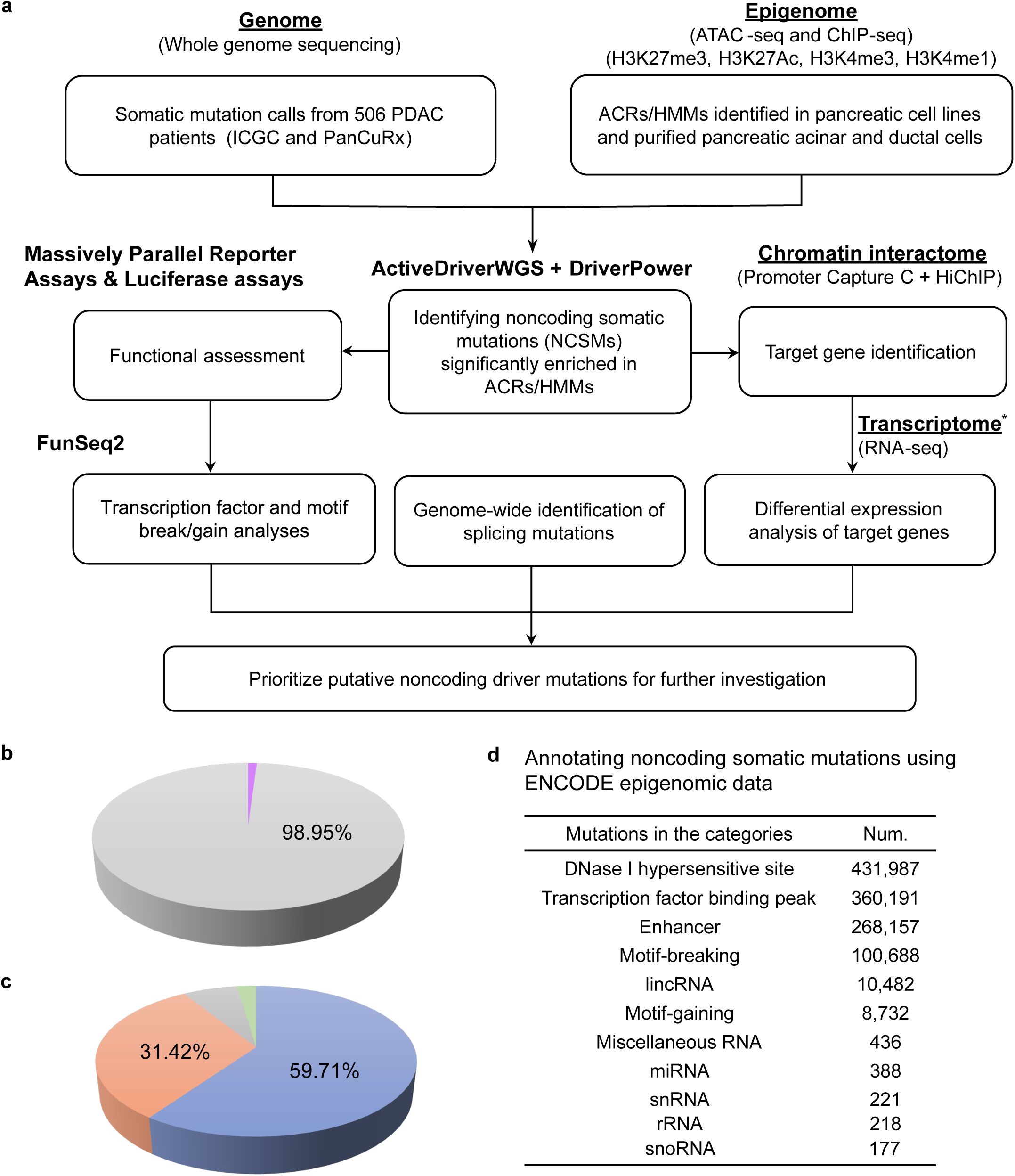
Study design and general annotations for somatic mutations identified in 506 PDAC patients. **a**, Study design for the integrated multiomic approach to identify noncoding driver mutations for PDAC used int his study. **b**, Somatic mutation calls from ICGC and PanCuRx in coding (purple, n = 36,490 or 1.05%) and noncoding (grey, n = 3,446,276 or 98.95%) regions of the genome (n = 506 PDAC patients after QC, see Methods). **c**, Classification of all NCSMs using ANNOVAR. Mutations are shown in intergenic regions (blue, n = 2,057,806 or 59.71% of the noncoding region), introns (orange, n = 1,082,746 or 31.42%), noncoding RNA introns (grey, n = 225,804 or 6.55%), Mutations in the remaining categories are shown as other (green) in the pie chart and consist of mutations in 3’UTRs (yellow, n = 26,493 or 0.77%), downstream of genes (n = 17,556 or 0.51%), upstream of genes (n = 17,389 or 0.50%), noncoding RNA exons (n = 11,818 or 0.34%), 5’UTR (n = 5,363 or 0.16%), both upstream and downstream of genes (n = 622 or 0.02%), splicing mutations (n = 603 or 0.02%), noncoding RNA splicing mutations (n = 49), both 5’UTR and 3’UTR (n = 23), and, both noncoding RNA exon and splicing mutations (n=4). **d**, Classification of NCSMs annotated by ENCODE epigenetic data from 147 cell lines and tissue samples. * Two RNA-seq datasets from the ICGC (n = 280) and PanCuRx (n = 246) consortia were used for differential expression analysis.

### Quality control and whole-genome annotation of somatic mutations from 506 PDAC patients

Whole genome somatic mutation calls from PDAC tumors were obtained from the International Cancer Genome Consortium (ICGC-PACA-AU and ICGC-PACA-CA)^16,17^ and the Pancreatic Cancer Translational Research Initiative (PanCuRx)^18^. Following quality control, samples from 506 patients remained (see Methods), with a mean somatic mutation count of 6,883 (range 102 – 46,628). The most frequently mutated protein coding genes across the three studies were similar, including *KRAS*, *TP53*, *CDKN2A* and *SMAD4* (Extended Data Fig. 1-3). The somatic mutation rate in the combined dataset averaged 2.22 mutations per Mb and most somatic mutations were in noncoding regions of the genome (98.95%) (Fig. 1b), in which 59.71%, 31.42% and 6.55% located to intergenic, coding intronic, and noncoding RNA intronic regions, respectively (Fig. 1c).

We first annotated somatic mutations using the FunSeq2 pipeline and ENCODE (The Encyclopedia of DNA Elements) data across all available cell types and tissue samples. We noted 431,987, 360,191 and 268,157 mutations in DNase I hypersensitive sites, transcription factor binding peaks and enhancers, respectively (Fig. 1d). Of these, 100,688 mutations were predicted to disrupt transcription factor (TF) motifs while 8,732 resulted in gained TF motifs. The top recurrently mutated non-protein-coding regions (annotated by ENCODE) (Supplementary Table 1) include enhancers at chr6:68,590,900-68,601,300 (mutations observed in 40 out of 506 PDAC patients; 7.91%) and chr17:57,913,400-57,927,100 (33 patients; 6.52%), and two lincRNAs, *TSIX* and *XIST* on chrX:73,012,040-73,049,066, (35 patients; 6.92%). The most frequently recurrent NCSMs (identified in ≥ 10 patients, or in 1.98-2.77% of patients, n = 24 NCSMs total) are listed in Supplementary Table 2.

### Mapping pancreatic accessible chromatin regions (ACRs) and histone modification marks (HMMs)

To identify enrichment of noncoding mutations in gene regulatory elements that are most relevant to normal- and tumor-derived pancreatic tissues, we performed ATAC-seq and histone ChIP-seq analysis in two normal-derived (HPDE-E6E7 and hTERT-HPNE) and six tumor-derived (COLO357, KP-4, PaTu8988T, SU.86.86, MIA PaCa-2, and PANC-1) pancreatic cell lines (Table 1). Moreover, we re-analyzed data generated in FACS-purified primary acinar (n = 3 for ATAC-seq, n = 2 for ChIP-seq) and ductal (n = 4 ATAC-seq, n = 3 ChIP-seq) cell populations obtained from donor pancreas with no reported pancreas dysfunction^22^. This identified 28,346-258,494 ACRs in primary pancreatic cell populations and cell lines (Table 1, Supplementary Tables 3 and 4, Extended Data Fig. 4 and 5, see Methods). Likewise, we mapped genomic regions with histone modifications typically enriched at promoters, enhancers, and repressed chromatin, including H3K4me1 marks (n = 73,249-267,643), H3K4me3 marks (n = 27,693-174,233), H3K27ac marks (n = 86,479-279,075), and H3K27me3 marks (n = 20,848-48,886) in the same cell lines and pancreatic exocrine cells (Table 1). We then used ChromHMM to model 8 chromatin states (weak enhancer 1, weak enhancer 2, active enhancer, active genic enhancer, active transcription start site (TSS), bivalent TSS, repressed, and quiescent; Extended Data Fig. 6)^23^ and ROSE2 to map super-enhancers (SE, n = 851-2,107) across the same pancreatic cell lines and sorted acinar and duct cell populations (Table 1). In total, we identified 2,929,610 ACR and HMM annotations that were used to assess the enrichment of somatic PDAC mutations.

**Table 1.** Pancreatic datasets and epigenetic annotations used for noncoding somatic mutation enrichment analysis. The number and length of mapped accessible chromatin regions (ACRs), histone modification marks (HMMs) and super-enhancers (SE) defined in primary pancreatic acinar cell populations, primary pancreatic duct cell populations, normal-derived pancreatic cell lines and tumor-derived pancreatic cell lines are shown. ^#^Noncoding gene regulatory elements from HPDE-E6E7 and hTERT-HPNE normal-derived pancreatic cell lines were merged prior to testing for enrichment of NCSMs. Likewise, elements from COLO357, KP-4, PaTu8988t, SU.86.86, MIA PaCa-2 and PANC-1 tumor-derived pancreatic cell lines were merged prior to the enrichment analysis. A total of 2,929,610 annotations (mapped in pancreatic acinar and ductal cell populations as well as in merged normal- and tumor-derived cell lines) were used for the noncoding somatic mutation enrichment analysis.

| Cell origin | Sample type | Feature | ATAC-Seq | H3K4me3 | H3K4me1 | H3K27ac | H3K27me3 | Super enhancer | Reference |
| --- | --- | --- | --- | --- | --- | --- | --- | --- | --- |
| **Purified pancreatic acinar and ductal cells** | Acinar cells | Number of elements | 47,943 | 36,316 | 130,407 | 86,479 | 40,792 | 1,830 | Arda et al., *Cell Systems*, 2018 |
|  |  | Average length (bp) | 338 | 382 | 395 | 453 | 11,642 | 24,128 |  |
|  |  | Total length (bp) | 16,197,723 | 13,873,134 | 51,573,090 | 39,211,311 | 474,885,708 | 44,154,250 |  |
|  | Ductal cells | Number of elements | 28,346 | 47,523 | 145,985 | 115,944 | 48,886 | 2,013 |  |
|  |  | Average length (bp) | 442 | 385 | 373 | 578 | 16,984 | 30,871 |  |
|  |  | Total length (bp) | 12,536,050 | 18,277,877 | 54,509,008 | 67,044,666 | 830,270,614 | 62,143,917 |  |
| **Normal-derived pancreatic cell lines** | HPDE-E6E7 | Number of elements | 109,212 | 127,685 | 209,871 | 165,634 | 32,138 | 1,304 | This study |
|  | hTERT-HPNE | Number of elements | 258,494 | 174,233 | 267,643 | 231,431 | 20,848 | 1,914 |  |
|  | Merged<sup>#</sup> | Number of elements | 110,559 | 202,545 | 341,357 | 289,299 | 33,325 | 2,469 |  |
|  |  | Average length (bp) | 570 | 860 | 942 | 1,103 | 30,520 | 110,821 |  |
|  |  | Total length (bp) | 63,013,361 | 174,253,717 | 321,463,948 | 319,185,292 | 1,017,080,175 | 273,617,734 |  |
| **Tumor-derived pancreatic cell lines** | COLO357 | Number of elements | 216,322 | 132,215 | 73,249 | 132,680 | 32,172 | 851 | This study |
|  | KP-4 | Number of elements | 107,999 | 100,428 | 131,689 | 174,005 | 30,956 | 1,062 |  |
|  | PaTu8988t | Number of elements | 132,485 | 99,058 | 143,163 | 184,355 | 24,594 | 1,457 |  |
|  | SU.86.86 | Number of elements | 190,546 | 152,434 | 144,334 | 163,037 | 26,973 | 1,367 |  |
|  | MIA PaCa-2 | Number of elements | 151,448 | 27,693 | 188,962 | 279,075 | 24,547 | 2,107 |  |
|  | PANC-1 | Number of elements | 116,498 | 73,582 | 180,761 | 165,910 | 24,876 | 1,219 |  |
|  | Merged<sup>#</sup> | Number of elements | 134,666 | 215,439 | 380,899 | 443,624 | 39,164 | 3,800 |  |
|  |  | Average length (bp) | 620 | 936 | 946 | 1,182 | 39,477 | 114,452 |  |
|  |  | Total length (bp) | 83,492,920 | 201,728,939 | 360,373,279 | 524,426,840 | 1,546,084,336 | 434,916,231 |  |

### Identification of noncoding somatic mutations (NCSMs) significantly enriched in ACRs/HMMs

To identify noncoding gene regulatory elements enriched with NCSMs as compared to the background mutational rate, we applied ActiveDriverWGS^24^ and DriverPower^25^ (functional adjustment by DANN^26^ and Eigen^27^) to the epigenetic annotations mapped in: 1) FACS purified pancreatic acinar cells, 2) FACS purified pancreatic duct cells, 3) normal-derived pancreatic cell lines, and 4) PDAC-derived cell lines. We identified a total of 314 ACRs/HMMs significantly enriched with somatic mutations (FDR ≤ 0.05) across these cell types (Table 2, Supplementary Table 5). Where ACRs/HMMs overlapped coding sequences, we observed significant enrichment of somatic mutations in known protein coding driver genes including *KRAS*, *TP53*, *CDKN2A* and *SMAD4* (Supplementary Table 6).

**Table 2.** Pancreatic gene regulatory elements significantly enriched with somatic mutations in PDAC. The number of accessible chromatin regions (ACRs) histone modification marks (HMMs) and super-enhancers (SE) identified in (**a**) purified primary pancreatic acinar cells, (**b**) primary ductal cells, (**c**) normal-derived pancreatic cell lines and (**d**) PDAC cells that were used for the NCSM enrichment analysis. Likewise, the number of gene regulatory elements identified as enriched for somatic mutations as per ActiveDriverWGS and DriverPower analysis are listed.

**a. ACRs/CREs identified in purified pancreatic acinar cells enriched for somatic mutations**
| Regulatory elements | ATAC-Seq | H3K4me3 | H3K4me1 | H3K27ac | H3K27me3 | Super-enhancer |
| --- | --- | --- | --- | --- | --- | --- |
| Number of elements tested | 47,943 | 36,316 | 130,407 | 86,479 | 40,792 | 1,830 |
| ActiveDriverWGS enriched elements | 2 | 2 | 13 | 17 | 16 | 2 |
| DriverPower enriched elements | 1 | 0 | 2 | 5 | 0 | 0 |

**b. ACRs/CREs identified in purified pancreatic ductal cells enriched for somatic mutations**
| Regulatory elements | ATAC-Seq | H3K4me3 | H3K4me1 | H3K27ac | H3K27me3 | Super enhancer |
| --- | --- | --- | --- | --- | --- | --- |
| Number of elements tested | 28,346 | 47,523 | 145,985 | 115,944 | 48,886 | 2,013 |
| ActiveDriverWGS enriched elements | 1 | 10 | 11 | 14 | 11 | 2 |
| DriverPower enriched elements | 0 | 4 | 2 | 1 | 0 | 0 |

**c. ACRs/CREs identified in normal derived pancreatic cell lines enriched for somatic mutations**
| Regulatory elements | ATAC-Seq | H3K4me3 | H3K4me1 | H3K27ac | H3K27me3 | Super enhancer |
| --- | --- | --- | --- | --- | --- | --- |
| Number of elements tested | 110,559 | 202,545 | 341,357 | 289,299 | 33,325 | 2,469 |
| ActiveDriverWGS enriched elements | 5 | 10 | 15 | 12 | 11 | 1 |
| DriverPower enriched elements | 3 | 4 | 5 | 6 | 0 | 0 |

**d. ACRs/CREs identified in PDAC cell lines enriched for somatic mutations**
| Regulatory elements | ATAC-Seq | H3K4me3 | H3K4me1 | H3K27ac | H3K27me3 | Super enhancer |
| --- | --- | --- | --- | --- | --- | --- |
| Number of elements tested | 134,666 | 215,439 | 380,899 | 443,624 | 39,164 | 3,800 |
| ActiveDriverWGS enriched elements | 1 | 18 | 25 | 55 | 32 | 0 |
| DriverPower enriched elements | 5 | 8 | 11 | 17 | 0 | 1 |

These 314 enriched ACRs/HMMs contained 4,612 simple somatic mutations (SSMs) across the 506 samples, 3,614 of which were unique NCSMs. Although a majority of these were only observed in a single patient, 988 NCSMs located to ACRs/HMMs where at least two PDAC patients had a mutation within a 31bp window (selected as the longest ENCODE transcription factor (TF) motif) (Supplementary Table 7), including 45 TF motif break or gain events (Supplementary Table 8). ENCODE annotations for the most recurrent NCSMs among the 314 elements are listed in Supplementary Table 9.

### NCSM-mediated alterations of gene regulatory activity

To functionally assess if the 988 NCSMs highlighted by our enrichment analysis alter gene regulatory activity, we performed MPRA in two human pancreatic cancer cell lines (PANC-1 and MIA PaCa-2) and a human embryonic kidney cell line (HEK293T). Following quality control (Methods, Extended Data Fig. 7), 184 out of 915 testable mutations (20.11%, see Methods) exerted significant differential transcriptional activity between mutant (MT) and wild-type (WT) alleles in PANC-1 cells, 118 (12.90%) in MIA PaCa-2 cells and 300 (32.79%) in HEK293T cells (FDR ≤ 0.05) (Fig. 2a-c). Subsequently, these “MPRA-significant” mutations were further deemed “MPRA-functional” when either the mutant or wild-type allele demonstrated an average RNA/DNA TPM ratio that diverged substantially from a null distribution of RNA/DNA TPM ratios derived from the activity of all scrambled control sequences. Using this approach, 87 (9.51%), 50 (5.46%) and 117 (12.79%) MPRA-functional noncoding somatic mutations were identified in PANC-1, MIA PaCa-2 and HEK293T cells, respectively (in total, 178 unique NCSMs (19.45% of those tested) were MPRA-functional in one or more cell line) (Fig. 2d, Supplementary Table 10).

**Fig. 2.**
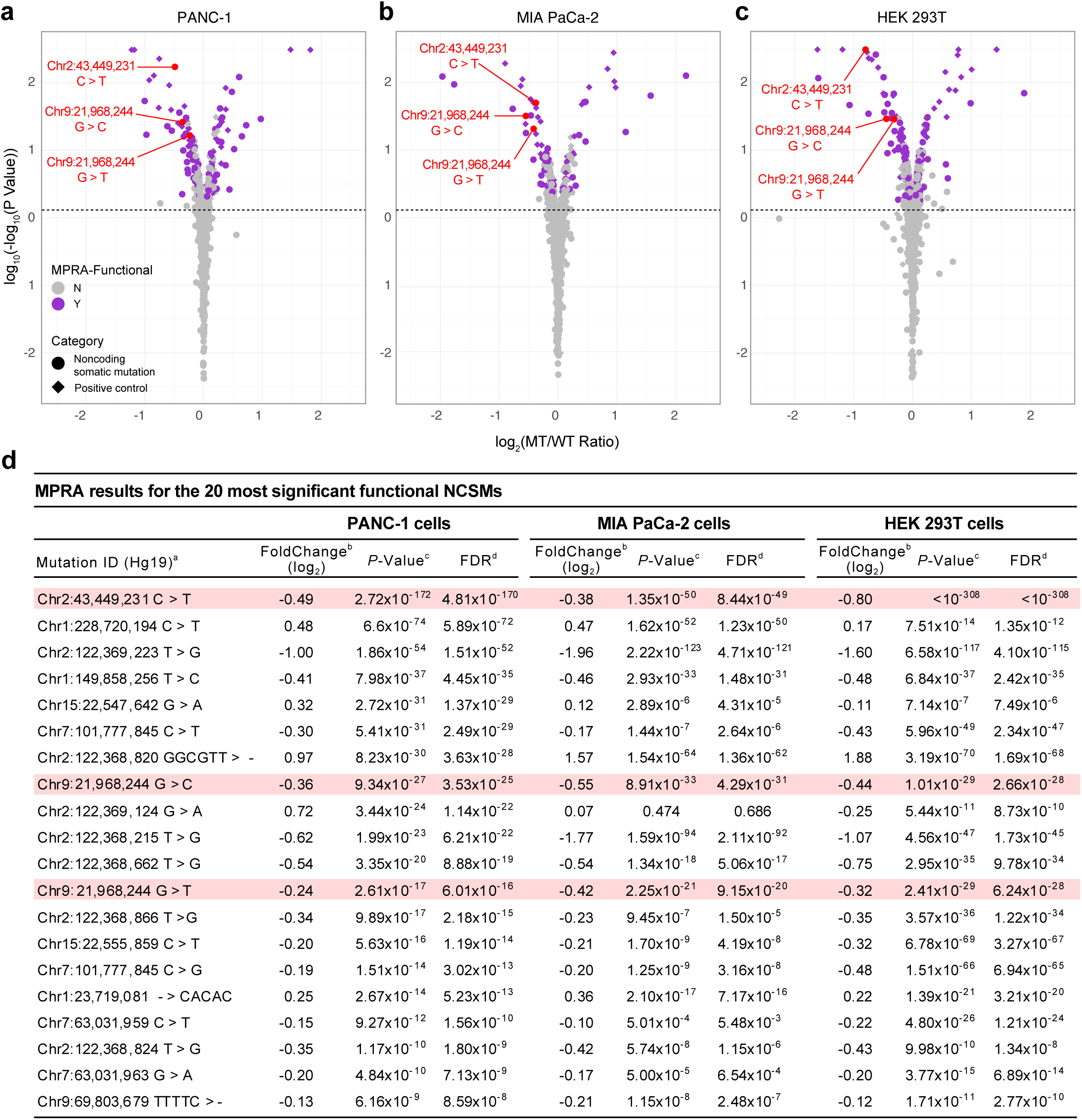
MPRA results for mutations enriched in accessible chromatin regions (ACRs) and histone modification marks (HMM) mapped in pancreatic cell lines and purified pancreatic acinar and duct cell populations. **a-c**, Volcano plots with MPRA results for PANC-1 (left), MIA PaCa-2 (center), and HEK293T (right) cells. MPRA effect sizes (Log2(MT RNA/DNA TPM)/(WT RNA/DNA TPM)) analyzed for the forward and reverse orientation inserts combined and significance (log10(-log10(*P*-value)) are show for all tested mutations. MPRA-functional mutations are shown as purple filled circles and all other mutations with grey filled circles on the plots in panels **a**, **b** and **c**. Mutations that were validated using individual luciferase assays are listed in red text with chromosomal location followed by the wild type and mutant alleles. **d**, MPRA results are listed for the 20 most significant NCSMs in PANC-1 cells, with corresponding results shown for MIA PaCa-2 and HEK293T cells. The three MPRA-functional mutations validated by individual luciferase assays are shown in pink highlights. MPRA regression results for all tested mutations and control variants are shown in Supplementary Table 10. Coordinates are based on human reference build hg19. MT: mutant allele, WT: wild type allele.

We selected the most significant MPRA-functional NCSM across the three cell lines (at chr2p21 near *ZFP36L2*, MPRA FDR-adjusted *P*-value rank 1, 7 and 1 in PANC-1, MIA PaCa-2 and HEK293T cells, respectively) and two mutations at chr9p21.3 in the last intron of *CDKN2A* (MPRA rank 10 - 18) for validation using individual luciferase assays (Fig. 3). The mutated allele (MT) at chr2p21 (310 bp downstream of the *ZFP36L2* gene, chr2:43,449,231 C>T), exhibited reduced gene regulatory activity in PANC-1 cells (MPRA log2(MT/WT) = -0.49, *P* = 4.81×10^-170^), MIA PaCa-2 cells (MPRA log2(MT/WT) = -0.38, *P* = 8.44×10^-49^) and HEK293T cells (MPRA log2(MT/WT) = -0.80, *P* < 1×10^-308^), which replicated by luciferase assays (PANC-1 cells: MT-T allele vs. WT-C allele fold change in the forward orientation = 0.36, *P* = 2.95×10^-7^; fold change in reverse orientation = 0.33, *P* = 3.42×10^-6^; MIA PaCa-2 cells: MT-T allele vs. WT-C allele fold change forward = 0.36, *P* = 3.99×10^-5^; reverse = 0.39, *P* = 8.73×10^-6^) (Fig. 3a and b). At chr9p21.3, two mutations were noted in an HMM in the last intron of *CDKN2A*. The former (chr9:21,968,244 G>T) was associated with reduced MPRA activity in PANC-1 cells (log_2_(MT/WT) = -0.24, *P* = 6.01×10^-16^), MIA PaCa-2 cells (log_2_(MT/WT) = -0.42, *P* = 9.15×10^-20^) and HEK293T cells (log_2_(MT/WT) = -0.32, *P* = 6.24×10^-28^). Lower activity was also observed for the latter mutation at the same genomic location (chr9:21,968,244 G>C) in PANC-1 cells (log_2_(MT/WT) = -0.36, *P* = 3.53×10^-25^), MIA PaCa-2 cells (log_2_(MT/WT) = -0.55, *P* = 4.29×10^-31^), and HEK293T cells (log_2_(MT/WT) = -0.44, *P* = 2.66×10^-28^). While significant differences were not observed in the forward orientation in individual luciferase assays, both the T and C mutations exhibited significantly (*P* ≤ 0.05, two-tailed unpaired T-test) lower relative luciferase values in the reverse orientation (PANC-1 cells: MT-T vs. WT-G allele fold change = 0.46, *P* = 1.22×10^-4^; MT-C vs. WT-G fold change = 0.22, *P* = 1.17×10^-5^; MIA PaCa-2 cells: MT-T vs. WT-G fold change = 0.59, *P* = 4.03×10^-7^; MT-C vs. WT-G fold change = 0.37, *P* = 2.66×10^-8^) (Fig. 3c and d).

**Fig. 3.**
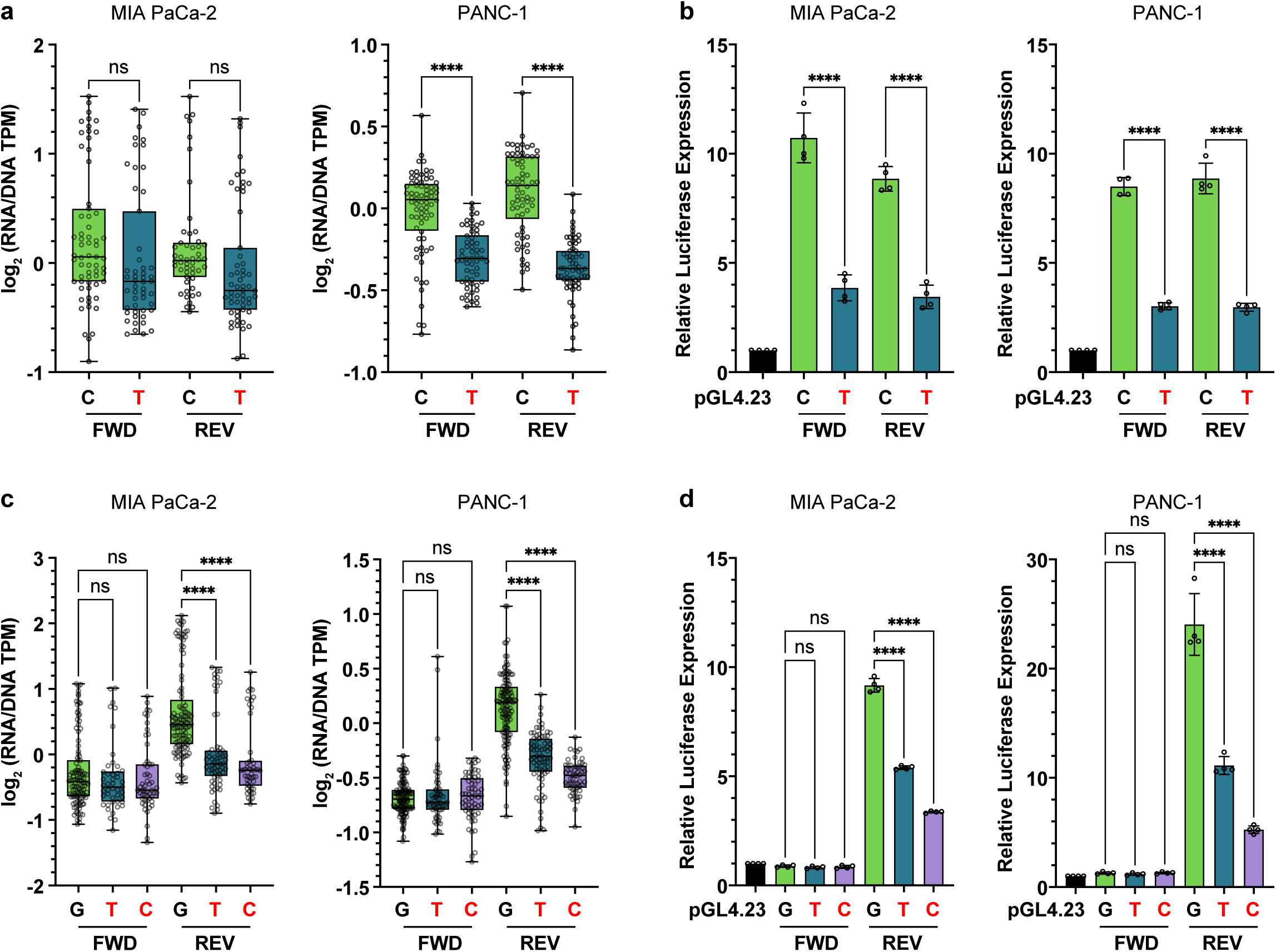
Luciferase validation for MPRA-functional NCSMs at chr2p21 near *ZFP36L2* and at chr9p21.3 intronic to *CDKN2A.* MPRA results (**a**) and luciferase validation (**b**) are shown for a NCSM at chr2:43,449,231 (C>T). MPRA results (**c**), and luciferase validation (**d**) for two mutations at chr9:21,968,244 (G>C and G>T). MPRA and luciferase results are shown for clones where the insert is in either the forward (FWD) or the reverse (REV) orientation. Mutated alleles are shown in red text. Stars indicate significance thresholds: \*\*\*\**P*-value < 0.0001; ns indicates nonsignificant results. Relative luciferase activity in **b** and **d** is compared to the empty vector. Error bars for the MPRA plots in panels **a** and **c** indicate the median, IQR (the boxes themselves) and the range of the data (whiskers) and for the luciferase assays in panels **b** and **d**, the S.E.M for four replicate experiments

### Identification of splicing mutations

We next investigated NCSMs in splice sites across the genome (using ANNOVAR annotations) and identified 361 patients (71.34%) with at least one splice site mutation considered damaging and possibly pathogenic. Of these, 30 patients (5.93%) had at least one recurrent mutation impacting splice sites in known tumor suppressor genes, including *TP53*, *CDKN2A,* and *SMAD4* (Supplementary Table 11). Splice site mutations in *CDKN2A* (chr9:21,968,244 G>T/C) and *SMAD4* (chr18:48,593,559 T>A/G), appeared to negatively affect splicing as indicated by RNA-seq reads showing disrupted normal splicing or skipping of specific exons (Fig. 4a, b, d, e; Extended Data Fig. 8a-b). Specifically, exon 2-3 splicing was disrupted in patients with *CDKN2A* splice acceptor mutations, and exon 10 skipping events were observed in patients with *SMAD4* splice site mutations (in frame). We confirmed the impact of predicted splicing mutations for both *CDKN2A* and *SMAD4* using exon trap assays in MIA PaCa-2 and PANC-1 cells (Fig. 4c, f; Extended Data Fig. 8c). The observed exon-skipping patterns were consistent with the predicted effects, suggesting a direct causal relationship. Notably, coding mutations in *CDKN2A* or in *SMAD4* were not observed in tumors from the PDAC patients harboring splicing mutations in these two genes. Of note, the two NCSMs in a splice acceptor site in the last intron of *CDKN2A* (chr9:21,968,244 G>T/C) negatively affected gene regulatory activity in both the MPRA and luciferase assays (as shown above).

**Fig. 4.**
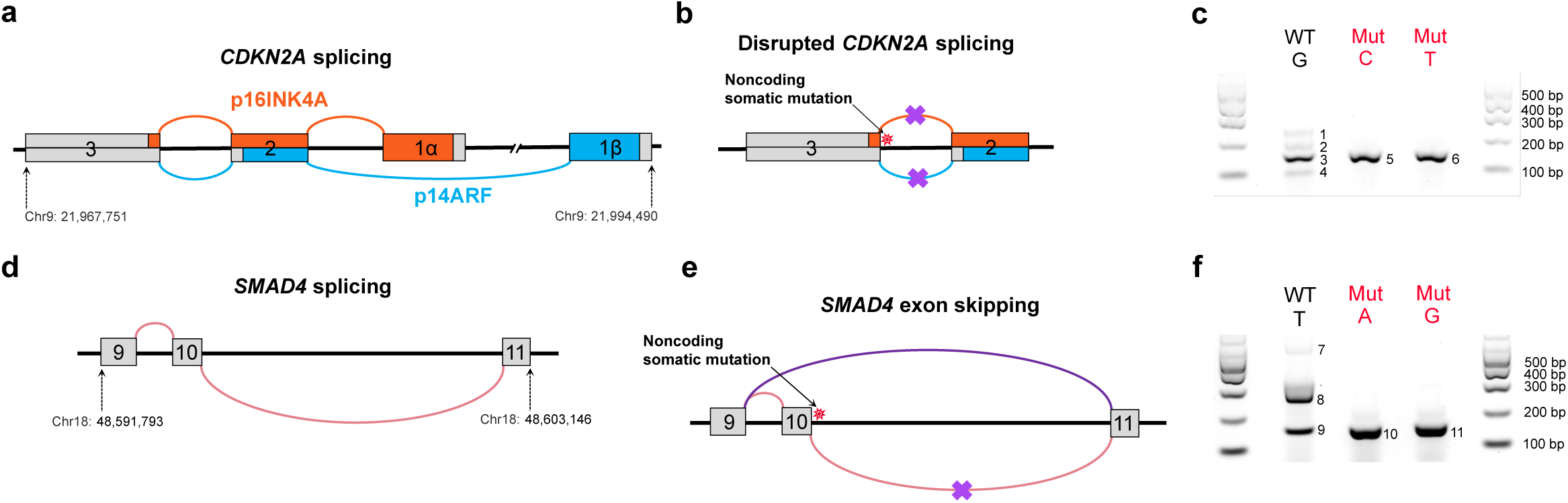
Functional assessment of splicing mutations in *CDKN2A* and *SMAD4.* **a**, A schematic figure showing *CDKN2A* exons 1 through 3 and the alternative splicing that generates both the p16INK4A (in red) and p14ARF (in blue) proteins. **b**, NCSMs at chr9:21,968,244 (G > C and G > T) in PDAC tumors from two patients (ICGC0536 and PCSI0777) are predicted to disrupt *CDKN2A* splicing between exons 2 and 3. **c**, Exon-trap validation for NCSMs at chr9:21,968,244 (G > C and G > T) in MIA PaCa-2 cells. **d**, A schematic figure of *SMAD4* exons and normal splicing between exons 9, 10 and 11. **e**, NCSMs at chr18:48,593,559 (T > A and T > G) are predicted to disrupt *SMAD4* splicing and result in exon 9 to exon 11 splicing thus skipping exon 10. These mutations were observed in PDAC tumors from two patients (PCSI0285 and PCSI0698). **f**, Exon-trap validation for the two NCSMs at chr18:48,593,559 (T > A and T > G) in MIA PaCa-2 cells (see Extended Data Fig. 8c for exon-trap validation in PANC-1 cells). Coordinates are based on human reference build hg19.

### Nominating candidate target genes for NCSMs

To identify putative target genes for mutations enriched in noncoding gene regulatory regions, we used proximity to transcriptional start sites and chromatin interaction analyses. Of the 988 NCSMs, 36 were directly located in promoters (see Methods) of 26 target genes (Fig. 5a and d) including *ZFP36L2* (encoding ZFP36 Ring Finger Protein Like 2, 6 mutations), *MIR21* (microRNA 21, 3 mutations) and *CPA3* (Carboxypeptidase A3, 1 mutation) (Supplementary Table 12). Subsequently, we generated chromatin interaction maps in MIA PaCa-2 and PANC-1 cells using high-resolution promoter-focused Capture-C (PCC)^20^, and observed 161 and 194 NCSMs in gene regulatory elements that physically interacted with the promoters of 166 and 183 potential target genes, respectively (Fig. 5b and e, Supplementary Table 13) including strong interactions (CHICAGO score ≥ 15, see Methods) with the promoters of *SNHG1* (small nucleolar RNA host gene 1) and *SLC3A2* (Amino acid transporter heavy chain SLC3A2) at chr11q12.13 and several core histone genes (*H2BC21*, *H2AC20*, *H2AC21*) at chr1q21.2. Likewise, using H3K27ac HiChIP-seq (HiChIP) data generated in purified pancreatic acinar and ductal cell populations, and in PANC-1 cells^28,29^, we identified 164, 137 and 122 NCSMs in regulatory elements that physically interacted with the promoters of 99, 124 and 107 target genes, respectively including strong interactions (PET counts ≥ 100, see Methods) with the promoters of *CPB1* (Carboxypeptidase B) and *AGTR1* (Type-1 angiotensin II receptor) at chr3q24, *MIR21* (microRNA 21) at chr17q23.1, and *SLC12A7* (Solute carrier family 12 member 7) at chr5p15.33 (Fig. 5c and f, Supplementary Tables 14 and 15). In summary, 416 potential target genes were identified, in which NCSMs localized to promoters or gene regulatory elements that physically interacted with gene promoters. Six target genes (*MTMR11, HIST2H2BC, BOLA1, ANP32E, SLC3A2* and *RALGDS*) were common among both the PCC and H3K27ac HiChIP-seq interaction data (Extended Data Fig. 9).

**Fig. 5.**
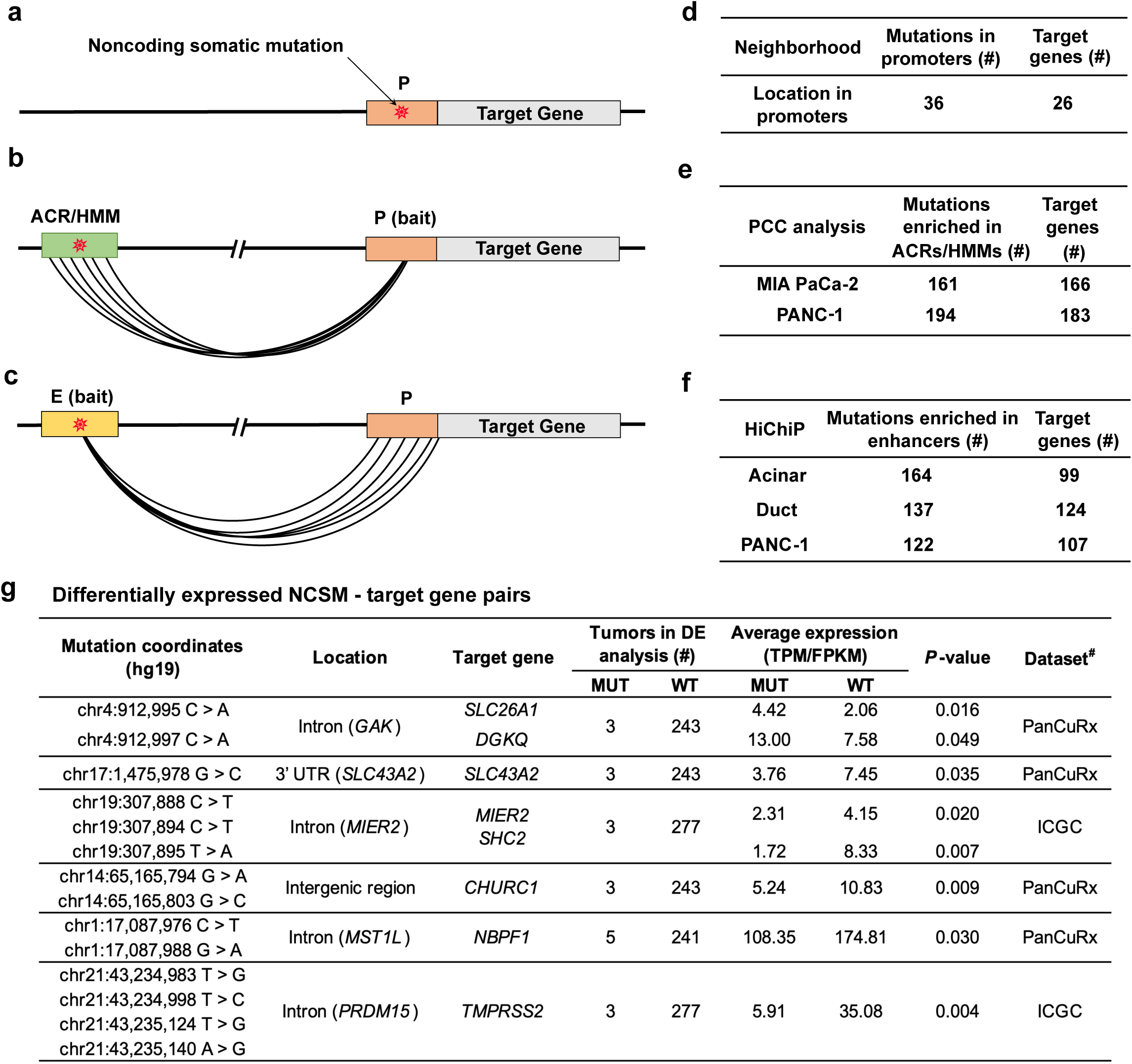
Identification of potential target genes for the 988 mutations enriched in pancreatic ACRs/HMMs. Schematic illustrations showing mutations falling: (**a**) directly within a promoter region, (**b),** in an ACR/HMM that physically interacts with a promoter region of a target gene (using chromatin interaction data detected by PCC in MIA PaCa-2 and PANC-1 cell lines), and, (**c**) in an enhancer region that physically interacts with a promoter region of a target gene (using chromatin interactions defined by H3K27ac HiChiP-seq in pancreatic acinar cell populations, pancreatic duct cell populations, and PANC-1 cells). **d,** Number of mutations enriched in promoters and their corresponding target genes. **e,** Number of mutations enriched in ACRs/HMMs and their target genes inferred by PCC. **f,** Number of mutations enriched in ACRs/HMMs, and their target genes inferred by H3K27ac HiChIP-seq. **g,** Differential expression analysis for PDAC samples with mutations as compared to nonmutated samples. This analysis was done separately for tumors from ICGC (n = 280) and PanCuRx (n = 246) with RNA-seq data. Significantly (Permutation test *P*-value ≤ 0.05) differentially expressed NCSMs - target genes. MUT: tumors with specific mutations. WT: tumors without the specific mutations. *Number of MUT and WT samples in the whole WGS dataset of 506 PDAC tumors. ^#^Dataset with significant differential findings (results from both ICGC and PanCuRx RNA-seq datasets for these genes is shown in Supplementary Table 16).

Using tumor-derived RNA-seq data from the ICGC (n=280) and PanCuRx (n=246), we tested if expression of each of the 416 target genes identified above differed significantly according to mutation status of their interacting gene regulatory element (see Methods). While statistical power is limited for this analysis, six significant NCSMs/target gene pairs (permutation test *P* ≤ 0.05) were observed (Fig. 5g) with: (i) higher expression of *SLC26A1* (fold change = 2.15, *P* = 0.016) and *DGKQ* (fold change = 1.72, *P =* 0.049) at chr4p16.3; (ii) lower expression of *SLC43A2* (fold change = 0.50, *P* = 0.035) at chr17:p13.3; (iii) lower expression of *MIER2* (fold change = 0.56, *P* = 0.020) and *SHC2* (fold change = 0.21, *P =* 0.0068) at chr19p13.3; (iv) lower expression of *CHURC1* (fold change = 0.48, *P* = 0.0095) at chr14q23.3; (v) lower expression *NBPF1* (fold change = 0.62, *P* = 0.030) at chr1p36.13; (vi) lower expression of *TMPRSS2* (fold change = 0.17, *P* = 0.0035) on chr21q22.3. However, none of these genes were significantly differentially expressed in both the ICGC and PanCuRx RNA-seq datasets (Supplementary Table 16).

### Identifying target genes for NCSMs at chr2p21 near *ZFP36L2*

The MPRA-functional, luciferase validated, NCSM at chr2p21 (chr2:43,449,231 C>T) localized to an ACR downstream of the *ZFP36L2* gene (Figure 6a), and to histone modification marks tagging promoters and enhancers. A nearby NCSM (chr2:43,449,213 - > AA) within this enriched ACR was also MPRA-functional in the three cell lines (log2(MT/WT) = -0.08 to -0.12, *P* (FDR corrected) = 0.01 to 5.4×10^-8^, MPRA rank 46 – 64 across the three tested cell lines, (Supplementary Table 10) but, due to a smaller effect size, was not validated with luciferase assays. Using the chromatin interaction datasets described above, we identified five potential target genes for this enhancer: *LINC01126, THADA, LINC02580, LINC01819* and *HAAO* (Fig. 6b). To identify genes regulated by this enhancer, we targeted it with CRISPR inhibition (CRISPRi) and measured the expression of these five genes, as well as that of *ZFP36L2* as the two mutations are only ∼4.5 kb away from its transcriptional start site, possibly limiting detection of chromatin interactions. Three of the four guides resulted in a significant reduction in *ZFP36L2* expression in PANC-1 cells (16.0 – 24.0% knockdown, *P*=0.005 - 0.023; the fourth guide showed a marginal reduction of 15.7%, *P* = 0.072) and one of four guides in MIA PaCa-2 cells (22.5% knockdown, *P* = 0.0081; three other guides showed reductions of 8.9 - 21.0%, *P* = 0.057-0.26) (Fig. 6c) indicating that it may represent a target gene for this enhancer. We also noted smaller reductions in one of the three tested *THADA* isoforms in PANC-1 cells (0 - 11.9%, *P* = 0.015 - 0.98) and MIA PaCa-2 cells (6.6% - 14.1%, *P* = 0.005-0.12) indicating that it may also be regulated by this enhancer. No effects were noted on expression of the other genes tested (Extended Data Fig. 10).

**Fig. 6.**
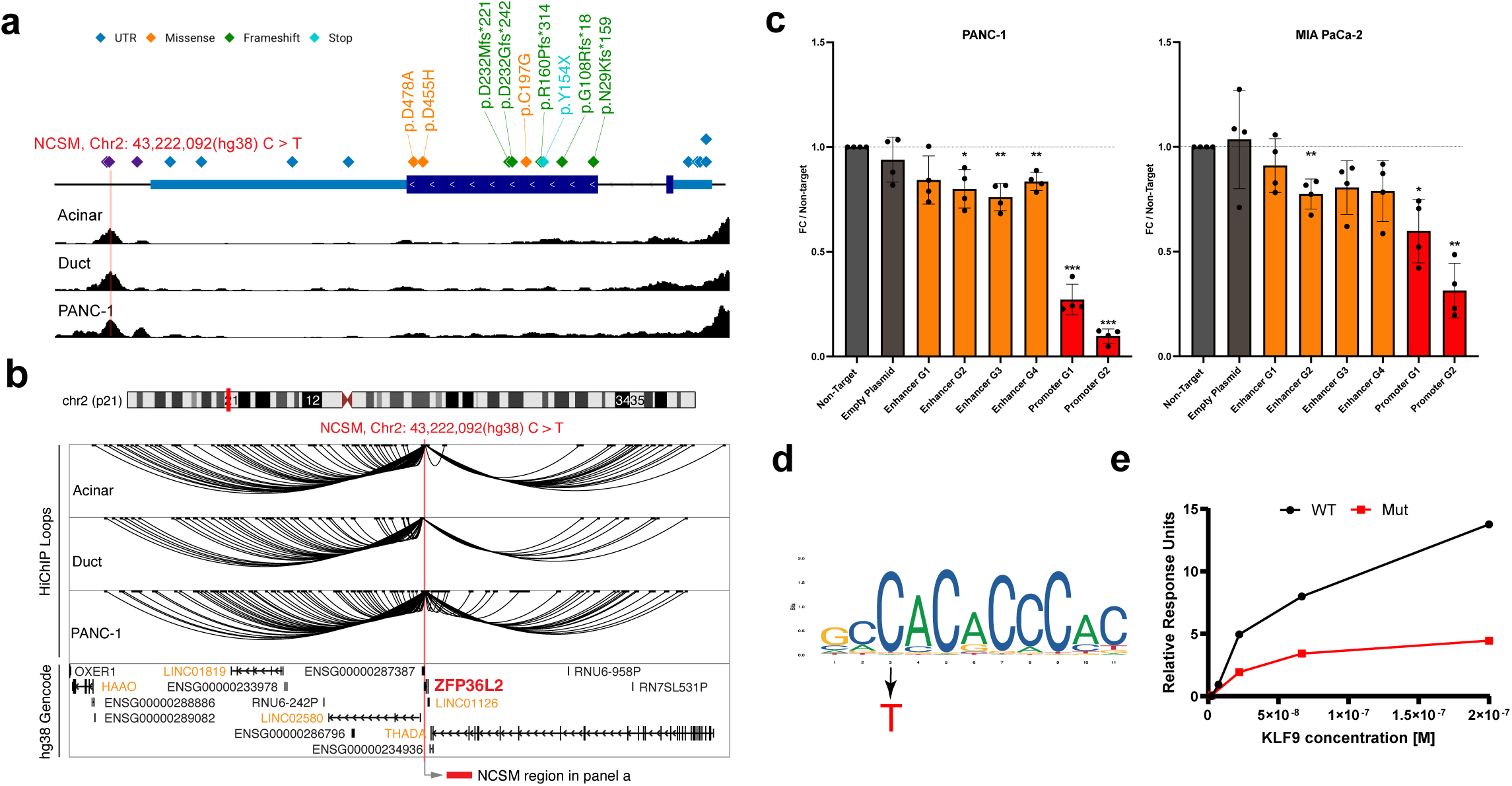
Potential noncoding driver mutations at chr2p21. **a**, A schematic illustration showing a NCSM (Hg38 chr2:43,222,092 C>T; Hg19 chr2:43,449,231) in an enhancer downstream of *ZFP36L2*. Additional mutations were located within the genes’ promoter, 5’UTR, exons, 3’UTR and the same downstream enhancer. The lower panel shows peaks of accessible chromatin (from ATAC-seq) in pancreatic acinar cells, pancreatic duct cells and PANC-1 cells (ATAC-seq results were similar in all other cell lines included in the study). **b**, Potential targets genes for the NCSM (chr2:43,222,092 C>T) assessed by H3K27ac HiChIP-seq in pancreatic acinar, ductal and PANC-1 cell lines. Interacting genes are listed in light orange text. **C,** CRISPRi targeting the mutated enhancer (dark orange) with four RNA guides as well as the promoter (red) of the gene with two guides in PANC-1 and MIA PaCa-2 cells. **d,** JASPAR DNA binding motif for KLF9 (MA1107.3) with the C>T mutation indicated (red). **e,** Surface Plasmon Resonance (SPR) results showing ∼3-fold lower binding of recombinant KLF9 protein to a oligo representing the mutation at chr2:43,449,231 (Hg19) and surrounding DNA sequence. Coordinates are based on human reference build hg19 except in panels **a** and **b**.

Mutations were also located directly in the 5’UTR/promoter (in 5 or 0.99% of all patients in our study), coding regions (in 9 patients; 1.78%) and the 3’UTR of *ZFP36L2* (4 patients; 0.79%). Coding mutations included frameshift (5 patients), nonsynonymous (3 patients) and stop-gain (1 patient) mutations indicating likely damaging effects on the protein. The mutation at chr2:43,449,231 C>T) is predicted to disrupt transcription factor binding motifs for multiple members of the Krüppel-Like (KLF) family of TFs, including KLF9 (Fig. 6d, Supplementary Table 17). Using Surface Plasmon Resonance (SPR), we validated this prediction and noted an approximately 3-fold reduced binding of recombinant KLF9 protein to the mutated (T) allele. Combined, our results indicate that KLF9, and possibly other members of this family of transcription factors, regulates expression of *ZFP36L2* and that somatic mutations at this site can lead to reduced expression of this gene (Fig. 6e).

## Discussion

The importance of somatic coding mutations in cancer initiation and progression has been well established since the discovery of a mutated *HRAS* oncogene over 40 years ago^30^. While most driver mutations have historically been identified in protein coding regions of the genome, noncoding somatic drivers are increasingly considered as important components of oncogenesis^31^. The vast size of the noncoding genome, combined with ambiguous effector mechanisms, complicates this search. Here, we present a comprehensive integrative analysis to nominate candidate noncoding somatic driver mutations for PDAC and identifying putative target genes. Our strategy employed three main steps. We first characterized the PDAC-relevant chromatin landscape by mapping ACRs and HMMs in tumor- and normal-derived pancreatic cell lines and purified pancreatic acinar and ductal cell populations, thus greatly expanding the breadth and scope of previously generated datasets for this organ. Subsequent enrichment analysis prioritized gene regulatory elements that may be somatically implicated in PDAC development. In the second step, we applied MPRA to identify functionally relevant somatic mutations in these elements, showing that a sizable proportion exerted significant disruptions to *cis*-regulatory activity. Finally, we applied an integrative approach to link mutated regulatory elements to putative target genes via chromatin interaction analysis and comparative transcriptomics, providing a valuable resource for understanding the downstream effects of noncoding mutations on gene regulation and possible mechanisms by which NCSMs may contribute to PDAC.

Our framework nominated mutations in an enhancer downstream of *ZFP36L2* as candidate noncoding drivers. These mutations decreased *cis*-regulatory activity for the surrounding gene regulatory element, that appears to drive *ZFP36L2* expression via KLF9. Additional mutations were located throughout the *ZFP36L2* gene in close to 4% of patients included in this study. The fact that most of the coding mutations were frameshift or stop-gain further supports a role for *ZFP36L2* as a pancreatic cancer driver gene with a possible tumor suppressor function. This gene encodes mRNA decay activator protein ZFP36L2, a zinc-finger RNA-binding protein of the tristetraprolin (TTP) family that regulates mRNA stability by binding and promoting the decay of adenylate- and uridylate-rich element (ARE)-containing mRNAs and plays important roles in controlling cell cycle progression, DNA damage repair, and immune responses^32,33^. However, one study has indicated that this gene may be overexpressed in PDAC where it drives cell proliferation and invasion *in vitro* supporting a possible oncogenic role^34^.

Three pan-cancer studies have investigated potentially functional noncoding driver mutations in 2,250^35^, 2,658^17^ and 3,949^15^ tumors by WGS, including PDAC. These studies used multi-tissue histone- and transcription factor annotations generated by ENCODE and the NIH Roadmap Epigenomics Consortia for enrichment analysis and nominated several noncoding mutations and target genes based on the closest genes. Coding mutations in *ZFP36L2* were identified in 0-8.3% of tumors across different organs (including 1.8 - 6.41% of PDAC) lending support to a possible tumor suppressor function. Noncoding driver mutations were not described, but promoter mutations were enriched in ovarian cancer (8.63%)^15^. Our results indicate that expression of this gene is also influenced by NCSMs in PDAC, in an enhancer downstream of the gene. Further work is warranted to investigate the role of *ZFP36L2* in PDAC development. Two studies have specifically focused on noncoding driver mutations for PDAC. The first assessed enrichment in ENCODE defined gene regulatory elements and then focused on mutations within gene promoters (+/- 2 kb of TSS)^36^. While noncoding mutations in promoters were identified, along with the pathways they may affect, specific mutations were not followed up with functional experiments. More recently, an analysis of CREs marked by H3K27ac in PDAC cell lines and patient-derived organoid samples was used to detect putative CRE drivers enriched for significant NCSMs, followed by STARR-seq reporter assays for candidate functional mutations; nominating 43 potential functional mutations^37^.

Beyond mutations that mediate their effect by disrupting active enhancer elements, we identified splicing mutations in *CDKN2A* and *SMAD4*, occurring in patients without coding mutations in each gene. Experimental validation indicated a direct causal relationship between these NCSMs and aberrant splicing, highlighting them as likely drivers that impact two well-known PDAC tumor suppressor genes. Surprisingly, mutations in a splice acceptor site in the last intron of *CDKN2A* influenced both splicing and gene regulatory activity. While effects of somatic mutations on splicing are already known at other splice sites in this gene, a possible gene regulatory effect at this genomic location requires further follow up to better define its function and target gene(s).

Our study provides important advantages. First, we focused our search for noncoding driver mutations on genome wide ACR and HMM maps derived from pancreatic-derived cell lines and primary pancreatic acinar and duct cell populations. As compared to previously available datasets with only small number of pancreatic cell lines and bulk pancreatic tissue samples, the maps we generated should represent a more refined picture of gene regulatory elements in the pancreas. Likewise, we used somatic mutation calls from the largest available WGS datasets generated for PDAC. Specifically, we used enrichment methods^25^ that outperform several other methods for noncoding somatic driver discovery and MPRA to start functionally assessing the effects of somatic noncoding mutations on gene regulatory activity. Furthermore, instead of using nearest genes to nominate target genes for enriched NCSMs, we conducted genome-scale chromatin interaction analysis from normal-derived pancreatic cell populations and PDAC cell lines to link mutations to target genes. Despite these strengths, our approach is not without its limitations. Foremost, given the inherent challenges of enrichment analyses for the mostly non-recurrent somatic noncoding mutations our work (and that of previous investigators) has identified, our analysis remains underpowered to characterize potentially important NCSMs in personal genomes that require even larger sample sizes. Likewise, differential expression analysis in tumors with and without specific mutations is underpowered due to the low frequency of most mutations and mutated gene regulatory elements. Furthermore, as the activity of noncoding regulatory elements depends on cellular context, developmental stage, and environmental stimuli, future studies incorporating such conditions are likely to enhance discovery of noncoding driver mutations. Simulating inflammation and ER stress might be important next steps for such an approach.

In conclusion, by creating maps of noncoding gene regulatory elements in pancreatic cell lines and purified exocrine cell populations, we have nominated and functionally assessed candidate driver mutations for PDAC and identified likely target genes. While noncoding mutational hotspots do not appear to be common in PDAC, it is likely that important gene regulatory elements are targets for mutations that may drive cancerous growth in this organ. Our findings contribute to understanding of the molecular mechanisms driving PDAC and provide avenues for further research with potential implications for patient care and outcomes for this cancer of dismal prognosis. Future follow up studies of the catalogs of candidate somatic driver mutations highlighted here, incorporated with noncoding germline risk variants^38^, epigenetic drivers^39^ and AI-powered approaches^40^, may therefore uncover more of the dark matter important for carcinogenesis in the noncoding genome.

## Online Methods

### Quality control and annotation of whole genome and RNA sequencing data

Whole genome sequence (WGS) data and somatic mutation calls from PDAC tumors arising in 787 patients were obtained from ICGC-PACA-AU (n=252), ICGC-PACA-CA (n=220)^16,17^ and PanCuRx (n=315)^18^. The details of these datasets were previously described^16–18^. To remove redundant SSMs for each patient, a single entry for each unique SSM was retained. To avoid outliers, tumors with less than 100 SSMs or a SSM count more than three standard deviations away from the mean were removed in each study (n=79, n=4 and n=5, respectively). Additionally, duplicate donors between ICGC-PACA-CA and PanCuRx were removed (n=193). In instances where tumor samples were sequenced multiple times, samples using laser capture micro-dissection (LCM) were retained. This resulted in data from a total of 506 PDAC patients. Oncoplots of the 10 most frequently mutated genes were generated by Maftools (v1.6.00)^41^. The average somatic mutation rate per Mb was calculated based on the size of human reference genome (total 3,101,788,170bp, https://www.ncbi.nlm.nih.gov/grc/human/data?asm=GRCh37.p13). Three types of annotations were performed for all mutations using ANNOVAR^42^ (v2019-10-24): (i) gene-based (whether a mutation is intronic, 3′untranslated region (UTR), 5′UTR, intergenic, etc.), (ii) region-based (whether a mutation overlaps with certain regions of interest) and (iii) filter-based (whether a mutation is present or absent in the interested datasets and report its frequency, such as 1000 Genomes Project). We used 1KGP.ALL.sites (201508), gnomAD (v2.1.1), HRC (v1.1), avsnp (v150), COSMIC (v91, v100), CADD (v1.3), DANN (2019), Eigen score (2019), dbNSFP (v3.5c), dbscSNV (1.1), GWAVA (20150623), ExAC (20151129) and ClinVar (20200316) databases to annotate all the mutations. To remove likely germline variants, mutations with minor allele frequency > 1% in the 1000 Genomes Project (1KGP, 5,008 haplotypes, Phase 3, Aug 2015)^43^, the Haplotype Reference Consortium (HRC, 64,976 haplotypes, v1.1)^44^ or the Genome Aggregation Database (gnomAD, 15,708 whole genomes and 125,748 exome sequences, v2.1.1)^45^ were removed.

FunSeq2 (v2-1.2)^46^ (http://funseq2.gersteinlab.org/) was used to identify mutations in known ENCODE annotations (dNase I hypersensitive sites (DHS), transcription factor binding peaks (TFP), transcription factor bound motifs in peak regions (TFM) motif breaks or gains, ncRNA genes, transcription factor highly occupied regions^47^, etc.) defined by ENCODE across 147 different tissue samples and cell lines^48^. PERFECTOS-APE (PrEdicting Regulatory Functional Effect by Approximate *P*-value Estimation, uses TFBS models from: HOCOMOCO, JASPAR, HT-SELEX, SwissRegulon and HOMER)^49^ and motifbreakR^50^ were also used to identify TF binding sites and visualize the motifs. All presented coordinates are based on human reference hg19 unless otherwise specified.

### Gene expression analysis

For LCM tumor samples sequenced using WGS by PanCuRx, gene expression was determined using whole transcriptome sequencing (RNA-seq) in 246 patients^18^. Normalized gene or transcript expression data (TPM, Transcripts Per Million) was obtained from the PanCuRx Data Access Committee, Ontario Institute for Cancer Research, Toronto, Canada. For tumor samples sequenced using WGS by ICGC, gene expression was called using RNA-seq in 280 patients. Normalized gene or transcript expression data (FPKM, Fragments Per Kilobase of transcript per Million) was downloaded from the ICGC data portal. For patients in ICGC with multiple tumors RNA-sequenced, the priority of selection criteria was as follows: (i) using samples from LCM primary tumor, (ii) LCM samples from metastatic tumors, (iii) LCM xenografts, (iv) non-LCM tumors. Genes or transcripts with > 0.1 TPM or FPKM in more than 3 patients were retained, which left 36,155 genes or 39,937 transcripts for further analyses in the PanCuRx and ICGC RNA-seq datasets respectively.

### Mapping accessible chromatin regions (ACRs) in pancreatic cell lines

ATAC-seq libraries were prepared in duplicate from biologically independent logarithmically growing cell cultures (harvested ad distinct passage numbers) for hTERT-HPNE (50k cells), HPDE-E6E7 (40k cells), PANC-1 (15k cells), MIA PaCa-2 (40k cells), COLO357 (30k cells), KP-4 (30k cells), PaTu8988T (50k cells), and SU.86.86 (40k cells) pancreatic cell lines. For each replicate, cell aliquots were subject to lysis using 10mM Tris-HCL pH 7.4, 10mM NaCl, 3mM MgCl_2_ and 0.1% IGEPAL CA-630. Sequencing adapters were added to open regions of the genome using the Illumina Tagment DNA TDE1 Enzyme and Buffer Kit (Illumina, Foster City, CA). Illumina sequencing libraries were generated using the Illumina Nextera DNA Sample Preparation kit (Illumina, Foster City, CA) with dual indexing and 11-14 cycles of PCR amplification. The 16 ATAC-seq libraries (2 replicates per cell line) were sequenced as 2×150bp paired-end reads on the HiSeq 2500 (v4 chemistry) with two libraries run per lane on a single flowcell. Read alignment (to hg19), QC, and accessible chromatin peak calling was performed using the Kundaje lab’s pipeline for ATAC-seq/DNase-seq (https://github.com/kundajelab/atac_dnase_pipelines) on the 16 cell line-derived libraries just described as well as three purified pancreatic acinar cell samples and four purified pancreatic duct cell samples generated by a previous study^22^. Final peaks used for accessible chromatin regions (ACRs) annotations were based on true replicates or pooled pseudo-replicate peaks with an IDR < 0.05 among the duplicate cell line samples or among all purified acinar (n=3) or duct (n=4) samples. The ATAC-seq QC metrics and ACR summary statistics are detailed in Supplementary Table 3. While duplicate read rates were somewhat higher than current ENCODE recommendations for many samples, only high confidence peaks with an IDR<0.05 were used in our annotation sets and those peak counts were in line with ENCODE recommendations for all cell lines. Read depth and peak counts were lower for the previously published purified acinar and duct cell samples. ACRs for normal-derived cell lines (hTERT-HPNE and HPDE-E6E7) and tumor-derived cell lines (PANC-1, MIA PaCa-2, COLO357, KP-4, PaTu8988T, and SU.86.86) were also combined with the mergePeaks command in HOMER^51^ to form the “All Normal” and “All PDAC” ACR annotations, respectively.

The ACR landscape of the individual replicates generated in the cell lines and sorted primary cells were compared by principal component analysis and hierarchical clustering by Pearson and Spearman correlation (Extended Data Fig. 4). First, a standardized set of ACRs was generated as the union of ATAC-seq peaks across all samples using the multiinter and merge functions of the BEDTools suite^52^. Aligned reads for each sample were then assigned to the combined set of ACRs using the featureCounts function of the Subread package^53^. ACR read counts were then normalized by feature length followed by read depth to yield a normalized openness metric analogous to TPM. Principal component analysis (PCA) was performed on the samples based on the normalized openness of all ACRs, the results of which were plotted using the ggplot2 package in R (Extended Data Fig. 4a). Finally, Pearson and Spearman correlations between the normalized openness values of all samples were determined. Samples were hierarchically clustered in heatmaps using R (Extended Data Fig. 4b and c). The most consistent clustering was observed among the cell-sorted primary acinar and duct cells, and among the PANC-1, MIA PaCa-2, KP-4, and PaTu8988T cell lines.

For comparison with ENCODE pancreas ACRs that are mapped to GRCh38, coordinates for ACRs used in this study were lifted over from GRCh37 to GRCh38 with the UCSC Genome Browser LiftOver webtool (https://genome.ucsc.edu/cgi-bin/hgLiftOver). To determine how many ACRs were unique or in common among our normal pancreas-derived cell lines, PDAC-derived cell lines, sorted acinar cells, sorted duct cells, and ENCODE bulk pancreas samples (ENCSR251POP, ENCSR705KEB, and ENCSR808ZMK), we used the mergePeaks command in HOMER to partition overlapping ACRs into every possible combination of sample types, which is summarized in Extended Data Fig. 5. Notably, of the 189,581 ACRs reported for these ENCODE bulk pancreas samples, 91,082 (48.0%) overlapped the ACRs used for this study while the ENCODE bulk pancreas samples missed 86,324 ACRs found in the cell lines and purified acinar and duct cells.

### Mapping histone modification marks (HMM) in pancreatic cell lines

Histone modification-specific chromatin immunoprecipitation (ChIP) was performed in duplicate on the same cell lines used for ATAC-seq described above using the Cell Signaling SimpleChIP Plus Enzymatic Chromatin IP Kit (Cell Signaling, Danvers, MA). Chromatin immunoprecipitation for histone marks H3K27Ac, H3K4me1, H3K4me3, and H3K27me3 were performed using the following antibodies, respectively: Abcam ab4729, Abcam ab8895, Abcam ab8580, and Cell Signaling #9733. ChIP-seq libraries were generated for each chromatin immunoprecipitation and a 2% chromatin input control for each replicate with the Illumina NextSeq 500 High Output v2.5 75 Cycles Single Read Kit (Illumina, Foster City, CA) with 4 multiplexed samples per run on an Illumina NextSeq 500. Reads were aligned to the human genome (hg19) by BWA^54^, then QC (cross-correlation analysis and IDR)^55–57^ and histone mark peak calling (with MACS2 or SICER)^58,59^ were performed using our in-house pipeline (https://github.com/eiserdr/histone-chipseq) on our cell line ChIP-seq data as well as two purified acinar cell samples and three purified duct cell samples generated by a previous study^22^. Final peaks used for histone mark annotations and subsequent super-enhancer and ChromHMM analyses were called from the pooled aligned reads for cell line replicates and matched purified pancreatic cell types (i.e., acinar [n = 2] and duct [n = 3]). The summary of samples’ read depths and final totals of histone ChIP-seq peaks are listed in Supplementary Table 4.

ChromHMM^23^ was used for hidden Markov modeling of 8-chromatin states based on the above described histone modification ChIP-seq. The hg19 genome-aligned ChIP-seq reads for the 8 cell lines, 2 acinar samples, and 3 duct samples were first binarized in 200bp bins using the BinarizeBam command followed by training of the 8-state model with the LearnModel command. The chromatin states were assigned functional annotations based on the histone modification enrichment patterns as shown in Extended Data Fig. 6. Super-enhancers (SEs) were identified based on H3K27Ac ChIP-seq signal enrichments using the Rank Ordering of Super-enhancers (ROSE2) algorithm^60,61^. In brief, the algorithm stitched together H3K27Ac peaks within a given distance of each other (12.5kb for all samples here analyzed) to form broad enhancer regions that were then ranked by H3K27Ac ChIP-seq signal enrichment. Enhancer regions above the geometrically defined inflection point were classified as super-enhancers. Final super-enhancer counts for each sample type are listed in Supplementary Table 4. Track files for viewing ATAC-seq peaks, ChromHMM chromatin states, and super enhancer annotations in a genome browser may be downloaded from https://github.com/hoskinsjw/Pancreas_epigenomics_2024. High-resolution promoter-focused Capture-C (PCC) was performed as previously described for PANC-1 and MIA PaCa-2 cells^62^.

Overlap of mutations found in intergenic regions introns, 5’UTRs, 3’UTRs, noncoding RNA exons and introns, and splicing mutations with ACRs/HMMs mapped in purified pancreatic acinar, ductal cells, normal-derived pancreatic cell lines, and PDAC-derived cell lines, is listed in Extended Data Fig. 11. Mutations that overlapped ACRs and different HMMs in acinar, duct and normal and tumor-derived cell lines are shown in Extended Data Fig. 12 and Extended Data Fig. 13.

### Mutation enrichment analysis

The ACRs, HMMs and super-enhancer elements defined above (2,929,610 elements mapped in pancreatic acinar cells, ductal cells, merged normal-derived pancreatic cell lines and merged tumor-derived pancreatic cell lines) were integrated with mutation calls from WGS data from 506 PDAC patients (Table 1, Fig. 1) using ActiveDriverWGS^24^ (v1.0.1) (based on R v4.0.0) and DriverPower^25^ (v1.0.2) (functional adjustment in DriverPower used DANN^26^ (June 2019) and Eigen^27^ (June 2019)). A Poisson generalized linear regression model was used to compare the mutational burden of test regulatory elements against the expected mutational burden of a background window (50 kbps up- and downstream of elements) in ActiveDriverWGS. Regulatory elements ≤50 kb were retained for further analysis. The ACRs and HMMs that we generated in pancreatic cell lines and exocrine cell populations were used as genomic features, and the background mutation rate was calculated by gradient boosting machine (GBM) model in DriverPower. Extended Data Fig. 14 shows feature importance for the background mutation rate. Regulatory elements with false discovery rate (FDR) ≤ 0.05 in the above enrichment analyses were retained and considered statistically significant. NCSMs were mapped to the significant ACRs/HMMs using BEDTools^52^ (v2.30.0). The blacklists of ENCODE^63^ (v3) based on hg19 and hg38 were used to identify whether NCSMs were in problematic regions of the two reference human genomes. Then sliding window analysis was performed to select the NCSMs occurring in at least 2 patients (NCSMs within 31bp windows between patients within the significant ACRs/HMMs). This window was selected based on the longest TF motif observed in ENCODE data.

### MPRA plasmid pool design & cloning

Two oligo libraries (NC1 and NC2 for noncoding library 1 and 2) were designed as described previously^64^. To quantify transcriptional activity associated with either MT or WT alleles of each NCSM, 145-base sequences centered on either allele (hg38) in the forward and reverse orientations were assigned to 35 12-base tag sequences using the processVCF function of the mpradesigntools R package^65^. Tag sequences were preselected for balanced GC content, minimal homopolymer runs, and reduced internal hairpin propensity from the publicly available FREE barcodes database^66^. Tags were additionally filtered out should a match be observed between a given tag and an entry in the miRbase database^67^. Tag sequences were separated from the experimental 145-bases by contiguous KpnI (GGTACC) and XbaI (TCTAGA) restriction endonuclease recognition sequences and flanked by binding sequences for PCR primers (structured oligo sequence: 5’-ACTGGCCGCTTCACTG-145 bases-GGTACCTCTAGA-12 base tag-AGATCGGAAGAGCGTCG-3’). Each tested NCSM was accompanied by a matched scrambled control in both forward and reverse directions, which took the respective reference construct and randomly scrambled the central 20-base sequence, with subsequent filtering removing any de novo restriction enzyme recognition sites. Both oligo pools also contained 72 positive variants identified from previous MPRAs (Supplementary Table 18) and luciferase assays which significant allelic transcriptional shifts. Of the positive controls, 43 (59.7%) were MPRA functional in one or more cell lines. The Pearson correlation for the positive control MPRA log2FoldChange was 0.67 (PANC-1 vs. HEK293T cells), 0.75 (MIA PaCa-2 vs HEL293T cells) and 0.91 (PANC-1 vs. MIAPaCa-2 cells). Of the 988 prioritized NCSMs, 73 mutations could not be incorporated into the final MPRA design due to liftover incompatibilities with mpradesigntools or the presence of multiple KpnI, XbaI or SfiI recognition sequences within the contextual sequence. Oligos were synthesized on the Sureprint oligo synthesis platform (Agilent, Santa Clara, CA) across two pools, each containing approximately 100,000 oligos (denoted NC1 and NC2).

To clone MPRA oligonucleotide pools into the delivery vector, 10 fmol of reconstituted oligonucleotide were first amplified via emulsion PCR (ePCR) using Herculase II fusion polymerase (Agilent, Santa Clara, CA) with 2 μM of primers providing SfiI enzyme sites (Supplementary Table 19) with the Micellula DNA Emulsion & Purification Kit (EURx/CHIMERx, Milwaukee, WI). Amplified oligo pools were ligated to Illumina adapter sequences using the KAPA HyperPrep kit (KAPA Biosystems, Wilmington, MA) and qualified using the KAPA qPCR assay before assaying for tag recovery using an Illumina MiSeq v3 kit, 150 bp, paired end reads (Illumina, San Diego, CA). Subsequently, ePCR products subject to digestion with SfiI (NEB) into the complementary site of the pMPRA1 vector (#49349, Addgene, Watertown, MA) with Electroligase (NEB) using a 1:25 molar ratio of vector:insert. The resultant plasmid was then digested with KpnI and XbaI (NEB) between the 145 bp element and the 12 bp tag sequence, facilitating the subsequent ligation of a luc2 open reading frame cloned from complementary restriction sites of the pMPRA2 vector (#49350, Addgene, Watertown, MA). The resultant ligation product was electro-transformed into ONESHOT electrocompetent cells (Thermo Fisher Scientific, Waltham, MA) so that each construct was distinctly transformed at least ten times. The final cloned library, intended for transfection, was confirmed as a single band on an agarose gel after digestion with KpnI.

### MPRA transfection, cell culture and sequencing library generation

MPRA was performed in PANC-1, MIA PaCa-2 pancreatic cancer cell lines, and in HEK293T. Cells were seeded at 5 million cells per 10 cm^2^ dish, with sufficient dishes transfected to ensure each designed MPRA construct was transfected approximately 100 times per replicate. MPRA plasmid pools were transfected using 14 μg of plasmid DNA with Lipofectamine 3000 (Thermo Fisher Scientific, Waltham, MA). Transfections were performed across four biological replicates per cell line, with replicates being initiated from distinct cell passages. Total RNA was harvested 48 hours after transfection using the Qiagen RNeasy kit (Qiagen, Hilden Germany). Resultant total mRNA was isolated using the PolyA purist MAG kit (Thermo Fisher Scientific, Waltham, MA). Following isolation, mRNA was treated with Turbo DNase (Invitrogen). cDNA synthesis was performed using Superscript III with an oligo dT primer, as previously described. cDNA was subject to PCR using Q5 high-fidelity polymerase (NEB) using primers to incorporate Illumina TruSeq adapter sequences (Supplementary Table 19). Tag-seq libraries were also prepared using input DNA in the same manner. NC1 and NC2 pools for each cell line, along with the DNA input were sequenced on a single SP NovaSeq flow cell (100bp) single end reads (Supplementary Table 20).

### MPRA quality control and data analysis

Utilizing FASTQ files from transfected plasmid DNA or RNA transcript sequencing (Supplementary Table 20), we counted the number of reads which completely matched each 12 bp tag sequence in addition to downstream sequences inclusive of an XbaI recognition site and 20 bp of 3’ luc2 sequence. For each transfection, tag counts per million sequencing reads (TPM) were derived by dividing each raw tag count by the total number of sequence-matching tag counts per million reads. To infer the transcriptional activity attributable to a given element, a singular pseudo count was added to each TPM value, with a TPM then being derived by dividing each tag’s RNA TPM by its respective DNA count. For each transfection, the median ratio for all tags was calculated, and adjusted ratios were derived by dividing each individual ratio by its corresponding median to ensure standardized scaling across replicates. Finally, adjusted ratios were log2 transformed to infer an activity level associated with each tested tag sequence.

Across the three cell lines at least 97.05% of barcode sequences were detected in both input DNA libraries and RNA samples. Reproducibility between transfections was evaluated using Pearson correlation. Following removal of a single outlier replicate for each cell line, high correlation was observed for transcriptional activity levels across transfection replicates in the three cell lines (median Pearson’s R = 0.71, 0.44 and 0.97 for constructs in pool NC1; Pearson’s R = 0.94, 0.79 and 0.94 for constructs tested in pool NC2 in PANC-1, MIA PaCa-2 and HEK293T cell respectively, n = 4 transfections per cell line; Extended Data Fig. 7). Amongst experimental plasmid constructs (those constructs which were not scrambled negative controls) derived from the common pool of positive control sequences, the median Pearson’s R was 0.74, 0.77 and 0.95 for PANC-1, MIA PaCa-2 and HEK293T, respectively, indicative of robust experimental reproducibility. Transfection replicate T3 for PANC-1 MPRA pool NC1 and T1 for PANC-1 cells in MPRA pool NC2 were determined to be outliers based on Pearson correlation and were removed from downstream analysis (Extended Data Fig. 7). To ensure accurate representation in the subsequent analysis, tags with TPM counts of less than two were excluded to prevent low-abundance tags from distorting the data, leaving 89.65% of designed tags remaining for subsequent analysis. To assess the impact of either MT of WT alleles on transcriptional activity, we used a standard linear regression model while adjusting for the effect of orientation (forward or reverse) as a binary covariate and biological replicate as a categorical covariate. That covariate was omitted from the model for a subset of NCSMs that were only synthesized in the forward orientation (Supplementary Table 10). The significance of each model was determined using the robust sandwich type variance estimate Wald test to account for potential heteroskedasticity in the measurement error using the R package Sandwich^68^. *P* values for each linear model were corrected for multiple testing using a Benjamini-Hochberg adjusted *P* value (FDR) of less than or equal to 0.05 as the threshold used to nominate variants with significant transcriptional effects between alleles and the scrambled controls. As an additional control to determine whether tested elements exhibited divergent activity relative to a transcriptionally inert control, the average log_2_(RNA/DNA TPM) ratio for each allele was compared against a distribution of log_2_(RNA/DNA TPM) ratios from all scrambled control constructs tested within that cell line. First, the interquartile scrambled activity range for each cell line was determined, with experimental alleles being nominated as putative activators should their mean log_2_ RNA/DNA TPM ratio be greater than Q3. Similarly, experimental alleles were nominated as putative suppressors should their normalized activity be less than Q1. Mutations that exerted an FDR < 0.05 allelic effect in addition to either allele being nominated as a putative activator or suppressor were defined as “MPRA-functional”.

### Validation for specific mutations using dual luciferase reporter assays

The genomic regions centered on MPRA-functional mutations chr2:43,449,231 C>T and chr9:21,968,244 G>T/C and were synthesized as 145 bp gBlocks (Integrated DNA Technologies, Coralville IA) in both forward and reverse orientation (Supplementary Table 19) and cloned into the multiple cloning site of the pGL4.23[luc2/minP] (Promega, Madison, WI) luciferase vector. Plasmid inserts were Sanger sequence-verified to contain the correct inserts and mutations. The pGL4.23 Firefly reporter plasmid (and a Renilla luciferase control vector) were co-transfected into PANC-1 and MIA PaCa-2 pancreatic cancer cell lines at ∼70% confluency using Lipofectamine 2000 (Thermo Fisher Scientific, Waltham, MA). Luciferase activity was measured 48 hours post-transfection with the Dual Luciferase Reporter Assay System (Promega, Madison, WI). Firefly luciferase activity relative to Renilla luciferase activity was quantified on a GloMax Explorer microplate reader (Promega) and plotted as compared to the empty pGL4.23 luciferase vector. Triplicate experimental results were pooled and were subject to a two-sided unpaired two-sided T-test was applied to assess significance for allelic differences in luciferase activity using GraphPad Prism v10.

### CRISPRi targeting of a mutated enhancer at chr2p21 near *ZFP36L2*

CRISPRi guides were selected from the ‘CRISPR Targets’ UCSC genome browser(hg38) track to target the mutated enhancer downstream of *ZFP36L2* (enhancer mutations chr2:43,222,092 C>T and chr2:43,222,074 ->AA) (Supplementary Table 19). Homology arms were added to guides for cloning DNA inserts into the 5′BstXI-BlpI3′ digestion site of the pU6-sgRNA EF1Alpha-puro-T2A-BFP expression plasmid (#60955, Addgene Watertown, MA). As previously described^69^, gRNA oligos were phosphorylated and annealed before cloning to the plasmid backbone. Inserts were verified with Sanger sequencing.

To generate gRNA lentivirus, 4M HEK293T cells were plated in 10cm^2^ dishes 24-hours prior to transfection, after which 9ug of each gRNA, 4ug of packing plasmid psPAX.2 (Addgene, Watertown, MA) and 2ug of the envelope vector pMD2.G (#12259, Addgene, Watertown, MA) were diluted in OptiMEM medium and Lipofectamine 3000 (Thermo Fisher Scientific, Waltham, MA) according to manufacturer’s instructions. Media was refreshed 6-8 hours post-transfection and subsequent lentiviral particles collected 48-hours post transfection. Lentiviral particles were centrifuged, filtered and precipitated with polyethylene glycol (2X) overnight and concentrated by centrifugation the following day.

PANC-1 and MIA PaCa-2 cell lines were stably transduced with dCas9-KRAB-ZIM3 lentiviral particles (plasmid #154472, Addgene, Watertown, MA) using polybrene and selected with 10 μg/mL blasticidin. Clonal cell lines were established by limiting dilution, expanded, and confirmed for dCas9-KRAB-ZIM3 expression by Western blot. To transduce dCas9-KRAB-ZIM3 pancreas cells, PANC-1 and MIA PaCa2 cells were plated 24-hours prior to transduction. Concentrated virus particles were added to individual wells for each guide supplemented with 8μg/ml of polybrene. Media was changed 24-hours post transduction and supplemented with 8μg/ml puromycin for selection. Selection media was changed every 24-hours and cells harvested for RNA once all kill-control cells were dead (∼72 hours post transduction). Total RNA was extracted using a QIAGEN QIAcube connect instrument with the RNeasy mini kit (Qiagen, Hilden Germany). Total RNA (1μg) was used for cDNA synthesis with SuperScript III reverse transcriptase (Thermo Fisher Scientific, Waltham, MA). Real Time PCR was then performed using a Quantstudio 7 Flex system with the following Taqman assays (Thermo Fisher): *ZFP36L2* (Hs00272828_m1), *HAAO* (Hs00895710_m1), *LINC01126* (Hs03676090_s1), *THADA* (Hs00965192_m1, Hs00965183_m1, Hs00965181_m1). Relative expression changes were presented after normalization to each endogenous *HPRT1* (VIC probe) control and non-targeting plasmid control. Samples were run in 4 independent experiments. An unpaired t-test with Welch’s correction was used to test for statistical significance.

### Surface Plasmon Resonance analysis

Interactions between recombinant human KLF9 protein (TP760077, Origene, Rockville, MD), and double stranded DNA were analyzed by surface plasmon resonance using a Biacore T200 system (GE Healthcare). Mutant and wild type oligonucleotides overlapping the mutation at Chr2:43,449,231 C>T near *ZFP36L2* (Supplementary Table 19) (IDT, Coralville, IA) were annealed at 95°C for 10 minutes and cooled slowly to room temperature. A C1 chip (Cytiva, Marlborough, MA) was utilized to capture ∼2000 RU Neutravadin on all 4 flow cells followed by WT DNA on FC2 and Mutant DNA on FC4. KLF9 was prepared in degassed, filtered HBS-P+ (Cytiva, Marlborough, MA) with 20 uM ZnSO4. Experiments in high performance mode were carried out using five injections (30 uL/min) of increasing concentration of recombinant KLF9 protein (2.47-200 nM) passed over the sensor chip for 300 s followed by a 600 s dissociation. Following buffer and reference subtraction, data was evaluated utilizing the Biacore T200 evaluation software (GE Healthcare, Chicago, IL).

### High-resolution promoter-focused Capture C analysis

The high-resolution promoter-focused Capture C design was described previously^20^. DpnII restriction fragments containing promoters for coding mRNA, noncoding RNA, antisense RNA, snRNA, miRNA, snoRNA, and lincRNA transcripts (UCSC lincRNA transcripts and sno/miRNA used human reference genome hg19 annotations) totaling 36,691 RNA baited fragments throughout the genome^20^. Paired-end sequence reads from three biological replicates for the PANC-1 and MIA PaCa-2 tumor-derived pancreatic cell lines were preprocessed using the HICUP pipeline (v0.8.0)^70^ (reads were mapped using Bowtie2). Significant promoter interactions (score > 5) at 1-DpnII fragment resolution were called using CHiCAGO (v1.13.1) with default parameters except for minFragLen set to 75 and binsize set to 2500. Significant interactions at 4-DpnII fragment resolution were also called using CHiCAGO with artificial.baitmap and.rmap files in which DpnII fragments were concatenated in silico into 4 consecutive fragments^20^ using default parameters except for minFragLen set to 75, binsize set to 10,000, and removeAdjacent set to FALSE. NCSMs within significant ACRs/HMMs were mapped to promoter-interacting-regions (PIR) using BEDTools^52^ (v2.30.0).

### H3K27ac HiChIP-seq analysis

The HiChIP-seq analysis has been previously described^28,29^. Thirteen H3K27ac-HiChIP libraries (four samples each from acinar and six of duct cells purified from human donor pancreas tissues, and three from PANC-1 cells) were sequenced with 2 x 75bp runs on an Illumina NextSeq instrument to obtain more than 180 million reads per sample. Paired-end reads from these samples were aligned to hg38 using the Hi-C Pro pipeline^71^ with default settings. Hi-C pro trims, aligns, and assigns reads to MboI restriction fragments, filters for valid interactions and generates binned interaction matrices. We used two computational programs to call loops on the Hi-C pro processed samples: hichipper^72^ and FitHiChIP^73^. For hichipper, we used the peak calling options EACH, SELF which included each sample individually and only self-ligation reads. hichipper interactions were filtered based on a PET count ≥ 2 and a Mango FDR ≤ 0.01, which were classified as “significant loops”. For FitHiChIP, we used the “loose” parameter and a bin size of 5,000 kb to call loops at FDR ≤ 0.05. Loops were converted to bigBed format then uploaded to UCSC browser for visualization. To mitigate the variabilities across biological replicates, particularly in primary samples, we constructed a “reference” chromatin loop set for each cell type or cell line. We merged raw reads (fastq) from replicate samples based on cell type (acinar, duct, and PANC1). These merged reads were processed using Hi-C Pro, followed by loop calling using hichipper and FitHiChIP, as described above. We then intersected the hichipper and FitHiChIP loops for each cell type using BEDTools’ pairToPair function^52^ which identifies overlaps between loops with the same anchors. These common loops, identified by two independent loop callers, were considered “high confidence loops”. The resulting datasets were designated as “reference” chromatin loop sets for acinar and duct cells, as well as the PANC1 cell line.

### Differential expression analysis to identify potential target genes

Differential expression was assessed for each NCSM using permutation testing in samples from PanCuRx (n=246) and ICGC (n=280). For each NCSM/target gene pair, tumors were split into two groups: those with NCSM in the ACRs/HMMs (mutant, MUT) and those without NCSM in the ACRs/HMMs (wild type, WT). Using the normalized expression data described above for the target genes, the observed differential expression value (ODEV) was calculated by the mean expression of the patients with NCSM in the ACRs/HMMs minus the mean expression of the patients without NCSM in the ACRs/HMMs. The expression values for patients with NCSM and the expression values for patients without NCSM were permuted 100,000 times to generate 100,000 additional permuted differential expression values (PDEV) for randomization test. The empirical *P*-value was computed as the fraction of times (x/100,000) that a PDEV is stronger than the PDEV (when PDEV is positive) or weaker than the PDEV (when PDEV is negative). NCSM/target gene pairs with ≤ false discovery rate (FDR) 0.05 were considered statistically significant. NCSMs found in repeat regions (annotated by RepeatMasker (v3.0.1, RepBase library (2010-03-02)), last updated at UCSC genome browser (2022-10-18) (https://genome.ucsc.edu/cgi-bin/hgGateway) were removed.

### Identification and functional validation of splicing mutations

Splicing mutations were annotated by ANNOVAR^42^ (v2019-10-24) using dbscSNV^74^ (v11) which includes all potential human SNVs within splicing consensus regions (−3 to +8 at the 5’ splice site and −12 to +2 at the 3’ splice site). dbscSNV_ADA_SCORE and dbscSNV_RF_SCORE with > 0.6 were considered damaging. To functionally assess alternative splicing mutations in *SMAD4* and *CDKN2A*, genomic fragments surrounding the WT and MT genetic contexts of both mutations were synthesized as gBlocks (Integrated DNA Technologies, Coralville IA (Supplementary Table 19). Oligos encompassed upstream intronic sequences, the WT or MT splice site and the experimental exon followed by downstream intonic (*SMAD4*) or intergenic (*CDKN2A*) sequence. Sequences were flanked by XhoI and SacII restriction enzyme recognition sites to allow for cloning into the Exontrap vector pET01 (MoBiTec, Gottingen, Germany). Exontrap constructs were confirmed for the correct insert sequence using Sanger sequencing and transfected into PANC-1 and MIA PaCa-2 cells using Lipofectamine 2000 (Invitrogen) across 3 biological replicates for each cell line. 48-hours later, total RNA was extracted using the mirVana kit (ThermoFisher Scientific, Waltham, MA), followed by cDNA synthesis using SuperScript III reverse transcriptase in combination with the ExonTrap cDNA primer 01 (5’ - GATCCACGATGC - 3’). PCR was performed on cDNA using Exontrap primers 02 (5’ - GATGGATCCGCTTCCTGCCCC – 3’) and reverse primer 03 (5’ – CTCCCGGGCCACCTCCAGTGCC – 5’). Resultant PCR products were subject to gel electrophoresis on a 2% agarose gel stained with 1x ethidium bromide. Specific bands were gel extracted and subject to NanoPore sequencing (Oxford Nanopore Technologies) using the ligation sequencing kit with native barcoding addon, samples were normalized and multiplexed for sequencing on a single Flongle flow cell on an Oxford NanoPore GridIon instrument. Demultiplexed fastq files were aligned to a reference genome derived from the corresponding ExonTrap plasmid map and BAM files were visualized in IGV to confirm the presence or absence of experimentally spliced exons. Expected splicing aberrations for *CDKN2A* (lack of splicing between exons 2 and 3) and *SMAD4* (skipping of exon 9-10 and 10-11 splicing and direct splicing between exons 9-11) were confirmed with sequencing.

## Data availability

High-resolution promoter-focused Capture C data generated in tumor-derived pancreatic cell lines (PANC-1, MIA PaCa-2), ATAC-Seq and ChIP-Seq data from tumor-derived pancreatic cell lines (PANC-1, MIA PaCa-2, COLO357, KP-4, PaTu8988t and SU.86.86) and normal-derived pancreatic cell lines (HPDE-E6E7 and hTert-HPNE) have been deposited in SRA under accession number PRJNA1041452. The raw ATAC-Seq and ChIP-Seq sequencing data from purified pancreatic acinar and ductal cell populations were obtained from NCBI’s Gene Expression Omnibus (GSE79468). H3K27ac HiChIP-seq data from pancreatic acinar and duct cell populations is available through GEO (GSE245484). Raw whole genome and transcriptome sequence data for ICGC-PACA samples was obtained from The International Cancer Genome Consortium through controlled access (https://docs.icgc-argo.org). The whole genome somatic mutation calls and gene expression counts of ICGC-PACA samples were obtained from the International Cancer Genome Consortium (IGCG) data portal (https://dcc.icgc.org/) in July 2019 but now hosted at https://object.genomeinformatics.org (Bucket Name: icgc25k-open, a publicly available Object Storage Bucket, which uses the AWS S3 interface and is accessible using any S3 compatible object storage client). Raw whole genome and transcriptome sequence data, somatic mutation calls and gene expression counts from PanCuRx (European Genome-phenome Archive accession number EGAS00001002543) were kindly provided from the Ontario Institute for Cancer Research, Ontario, Canada (via controlled access).

## Acknowledgements

We would like to thank the Cancer Genomics Research Laboratory (CGR) of the Division of Cancer Epidemiology and Genetics (DCEG), National Cancer Institute (NCI), NIH, for sequencing and bioinformatics support. This study utilized the high-performance computational capabilities of the Biowulf Linux cluster at the NIH, Bethesda, MD, USA (http://biowulf.nih.gov). The authors would like to thank clinical contributors and data producers from the International Cancer Genome Consortium (ICGC) who have provided samples and data used in this work. This study was conducted using data provided by the Ontario Institute for Cancer Research which is funded by the Government of Ontario. The views expressed in the publication are those of the author and do not necessarily reflect those of the Ontario Institute for Cancer Research or the Government of Ontario.

## Funding

The work was supported by the Intramural Research Program (IRP) of the Division of Cancer Epidemiology and Genetics (DCEG), National Cancer Institute (NCI), US National Institutes of Health (NIH). The content of this publication does not necessarily reflect the views or policies of the US Department of Health and Human Services, nor does the mention of trade names, commercial products, or organizations imply endorsement by the US Government.

## Author Contributions

L.T.A and J.Z conceived the project. J.Z. designed and performed the major computational analysis and wrote the first draft of the manuscript. A.O.B performed MPRA, Luciferase validation and exon-trap experiments with help from I.C. and K.E.C. J.W.H designed and supervised ATAC-seq and ChIP-seq analysis. D.E and M.M performed ATAC-seq and ChIP-seq analysis. M.P performed CRISPRi experiments. H.E.A designed and supervised the H3K27ac HiChIP-seq analysis. L.W., K.G., and T.T. performed HiChIP analysis. M.O.N and T.A designed and preformed SPR analysis. C.D. and TZ helped with computational analysis. A.J., A.D.W., M.E.L, J.A.P., and S.F.A.G. generated PCC data. J.S. helped with the design of the enrichment analysis. J.Z, A.O.B, J.W.H and L.T.A finalized the manuscript with input from all authors.

## Competing Interests statement

The authors declare no competing interests.

## Supplementary information

**Supplementary Table 1:** FunSeq2 annotations for the most frequently mutated non-protein coding elements (in five ENCODE categories).

**Supplementary Table 2.** 24 Genome-wide identically recurrent (n ≥ 10) NCSMs in PDAC. Mutations were annotated by ANNOVAR.

**Supplementary Table 3**. ATAC-seq QC metrics and ACR summary statistics.

**Supplementary Table 4:** Histone ChIP-seq and super-enhancers summary statistics for normal-derived, PDAC-derived cell lines and purified acinar and duct cell populations.

**Supplementary Table 5:** All (n=314) pancreatic ACRs/HMMs significantly enriched with somatic mutations. A list of ACRs/HMMs that were significantly enriched with somatic mutations. Note that some elements overlapped between enrichment methods and were merged which resulted in a total of 314 ACRs/HMMs.

**Supplementary Table 6:** The top 15 ACRs/HMMs significantly enriched with somatic mutations. The ACRs/HMMs covering *KRAS, TP53, CDKN2A* and *SMAD4* had the highest tumor mutation burden.

**Supplementary Table 7:** List of the 988 NCSMs which were enriched in the pancreatic ACRs/HMMs. The coordinates are based on human reference builds hg19 and hg38, respectively. Three NCSMs in human reference hg19 could not be mapped to hg38 (marked unmap).

**Supplementary Table 8:** 45 motif break/gain events caused by the 988 NCSMs. The BED format details of FunSeq2 annotation can be accessed from: https://info.gersteinlab.org/Funseq2.

**Supplementary Table 9:** ENCODE annotations for noncoding mutations in the top significant pancreatic ACR/HMM (tumor- and normal-derived cell lines and acinar and duct cell populations). BED format details of FunSeq2 annotations can be accessed from: https://info.gersteinlab.org/Funseq2.

**Supplementary Table 10:** MPRA results for the 915 tested NCSMs and 72 positive controls. Linear regression results for each tested NCSM accompanied by log2(MT/WT (RNA TPM +1/DNA TPM +1)). *P* values and FDR-adjusted P-values for comparisons between MT and WT alleles in addition to comparisons between each tested allele and the average of all scramble sequences tested in that cell line.

**Supplementary Table 11:** Splicing mutations shared by ≥ 2 patients. All splicing mutations were with both dbscSNV_RF_SCORE and dbscSNV_ADA_SCORE ≥ 0.6 annotated by ANNOVAR using dbscSNV.

**Supplementary Table 12:** 36 NCSMs directly in gene promoters. Enriched NCSM that were in gene promoters as defined by HPCC baits.

**Supplementary Table 13:** Chromosomal interactions between the NCSMs and target regions by HPCC. The interaction data was analyzed by the ChICAGO pipeline.

**Supplementary Table 14:** Chromatin interactions between the NCSMs and target regions by HiChIP-seq (part 1, interactions to the right of mutation). The HiChIP data was analyzed using hichipper and FitHiChip.

**Supplementary Table 15:** Chromatin interactions between the NCSMs and target regions by HiChIP-seq (part 2, interactions to the left of each mutation). The HiChIP data were analyzed using hichipper and FitHiChip.

**Supplementary Table 16:** Genes with significantly differential expression (DE) in mutated tumors as compared to WT tumors in ICGC (n=280) and PanCuRx (n=246) samples with RNA-seq data

**Supplementary Table 17:** Transcription factor binding motif predication for a noncoding somatic mutation downstream of *ZFP36L2* (chr2: 43,449,231 C > T (hg19)). Prediction was performed using PERFECTOS-APE: PrEdicting Regulatory Functional Effect by Approximate *P*-value Estimation.

**Supplementary Table 18:** Positive controls used in MPRA experiments.

**Supplementary Table 19:** Experimental oligonucleotide/gBlock sequences.

**Supplementary Table 20:** Sequencing statistics for MPRA cDNA Libraries and Mapping of Tag Sequences.

## Notes

### Competing Interest Statement

The authors have declared no competing interest.

### Author Declarations

https://docs.icgc-argo.org https://ega-archive.org/studies/EGAS00001002543

### Summary of Updates

Manuscript pdf file now with text and figures of the same size. No changes were made in text or any content.

