## Extended Data Figures for "Large-scale multi-omic analysis identifies noncoding somatic driver mutations and nominates *ZFP36L2* as a driver gene for pancreatic ductal adenocarcinoma"

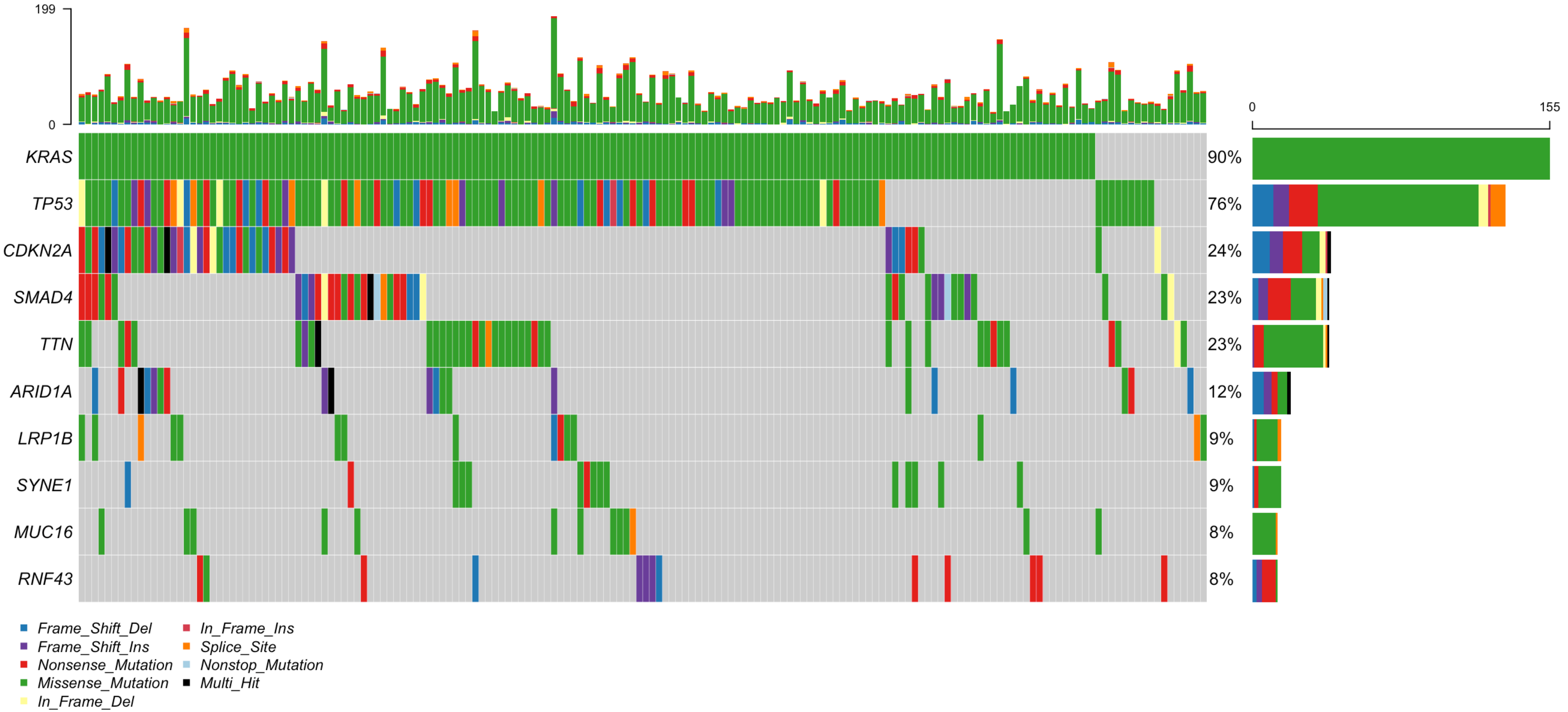


**Extended Data Fig. 1: Oncoplot of the top 10 most mutated genes in ICGC-PACA-AU (n = 173).** Genes were ordered by their mutation frequency. The percentages and the bar plots on the right represent the fraction and the number of patients with somatic mutations in the corresponding genes on the left. The types of mutation were color coded and listed at the bottom. All patients in this study with less than 100 SSMs or a SSM count more than three standard deviations away from the mean were removed. Mutations annotated as "Multi_Hit" were those genes which were mutated more than once in the same sample.

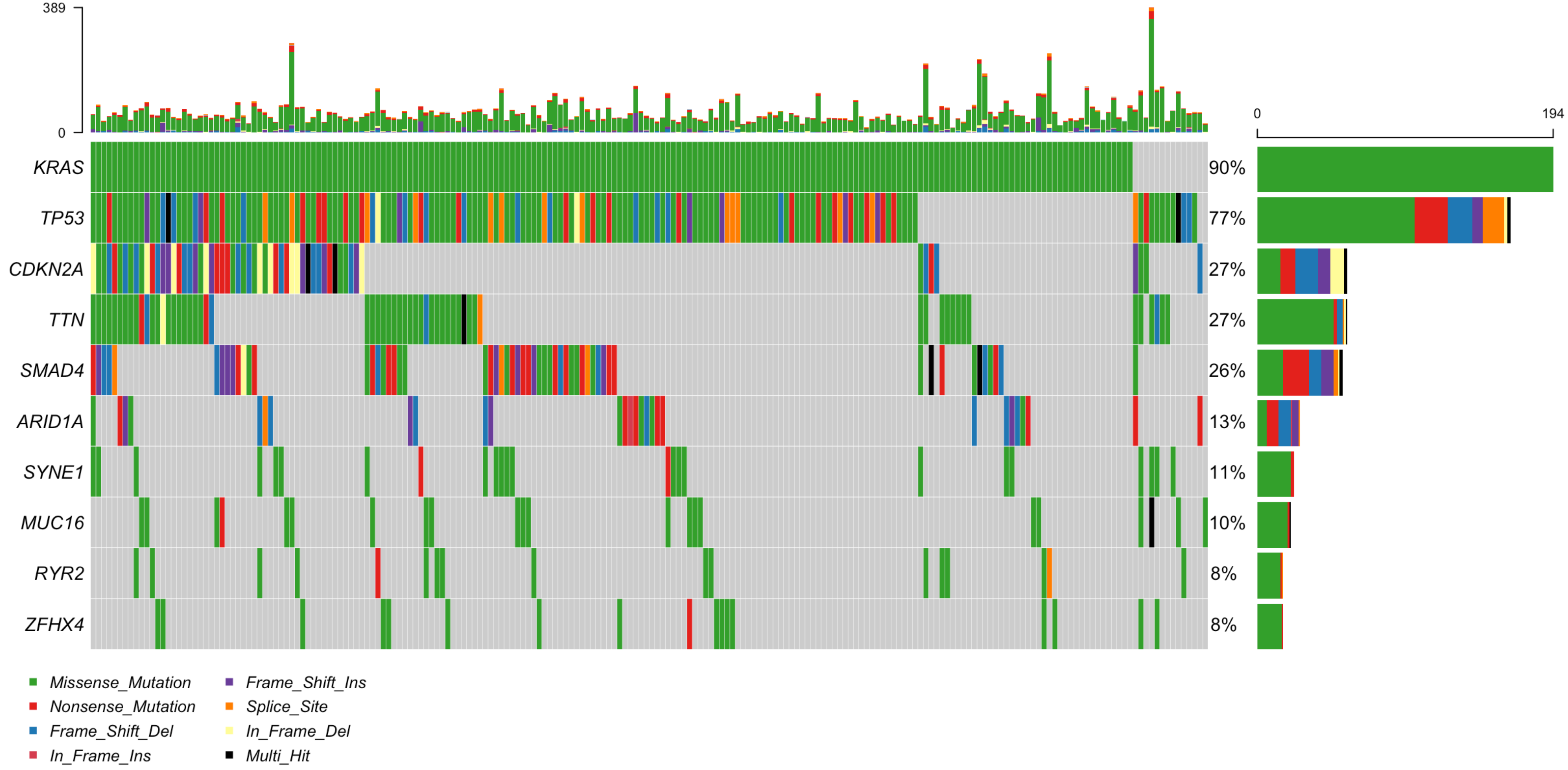


**Extended Data Fig. 2: Oncoplot of the top 10 most mutated genes in ICGC-PACA-CA (n = 216).** Genes were ordered by their mutation frequency. The percentages and the bar plots on the right represent the fraction and the number of patients with somatic mutations in the corresponding genes on the left. The types of mutation were color coded and listed at the bottom. All patients in this study with less than 100 SSMs or a SSM count more than three standard deviations away from the mean were removed. To observe the systematic pattern of mutated genes, the duplicate donors between the ICGC-PACA-CA and PanCuRx were included here. In the follow up mutation enrichment analysis, the duplicate donors between ICGC-PACA-CA and PanCuRx were removed.

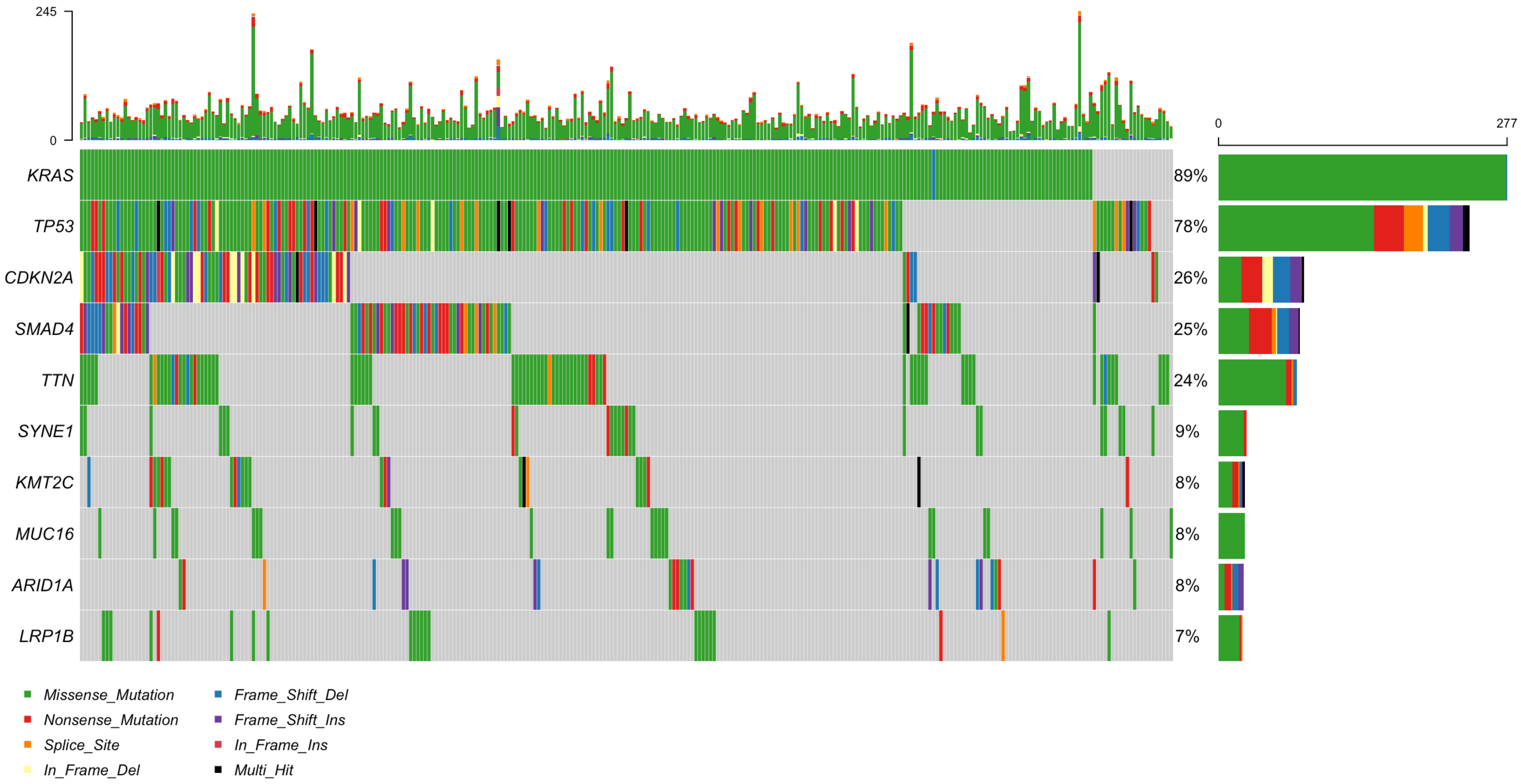


**Extended Data Fig. 3: Oncoplot of the top 10 most mutated genes in PanCuRx (n = 310).** Genes were ordered by their mutation frequency. The percentages and the bar plots on the right represent the fraction and the number of patients with somatic mutations in the corresponding genes on the left. The types of mutation were color coded and listed at the bottom. All patients in this study with less than 100 SSMs or a SSM count more than three standard deviations away from the mean were removed. To observe the systematic pattern of mutated genes, the duplicate donors between the PanCuRx and ICGC-PACA-CA were included here. In the follow up mutation enrichment analysis, the duplicate donors between PanCuRx and ICGC-PACA-CA were removed.

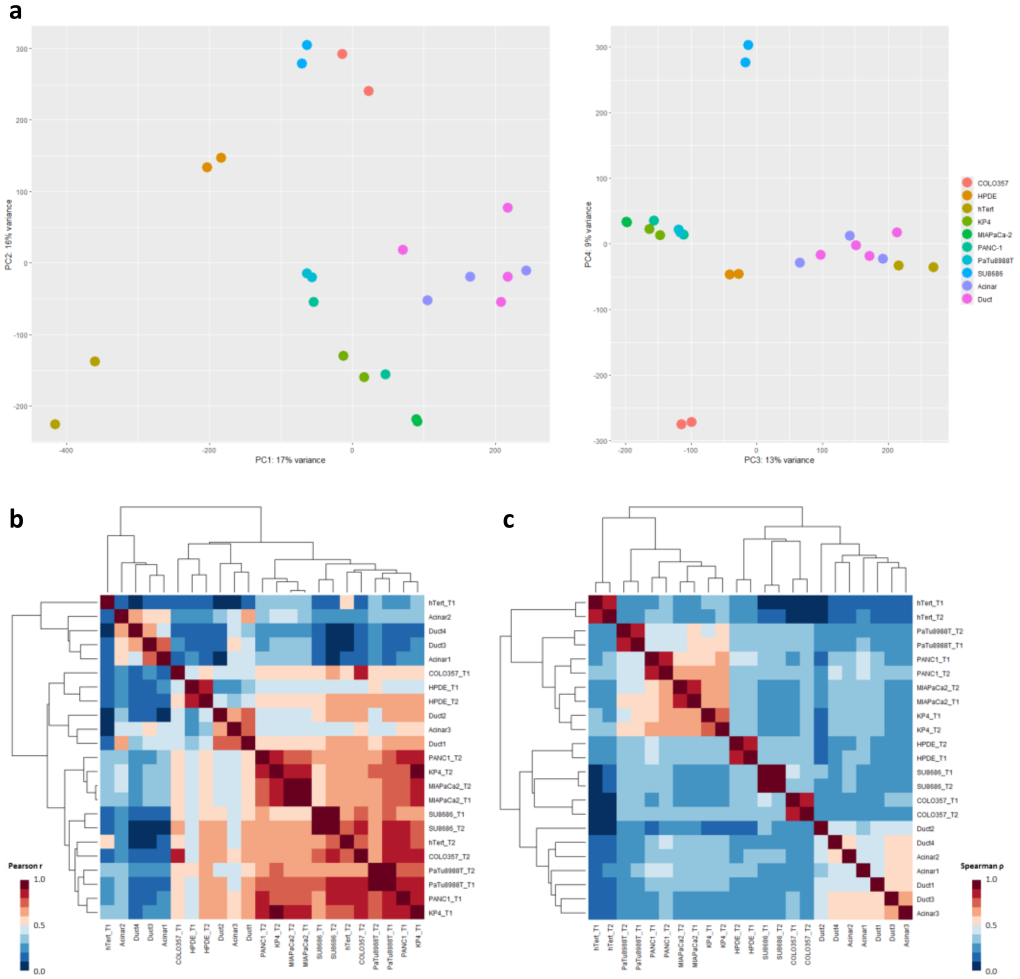


**Extended Data Fig. 4: ACR landscape comparisons of cell lines and sorted primary cell populations.** Scatter plots of PC1 vs. PC2 and PC3 vs. PC4 from the principal component analysis (PCA) of sample replicates based on normalized ATAC-seq read counts mapped to the union of all identified ACRs. (B and C) Hierarchically clustered heatmaps of Pearson (B) or Spearman (C) correlations between all sample replicates based on normalized ATAC-seq read counts mapped to the union of all identified ACRs.

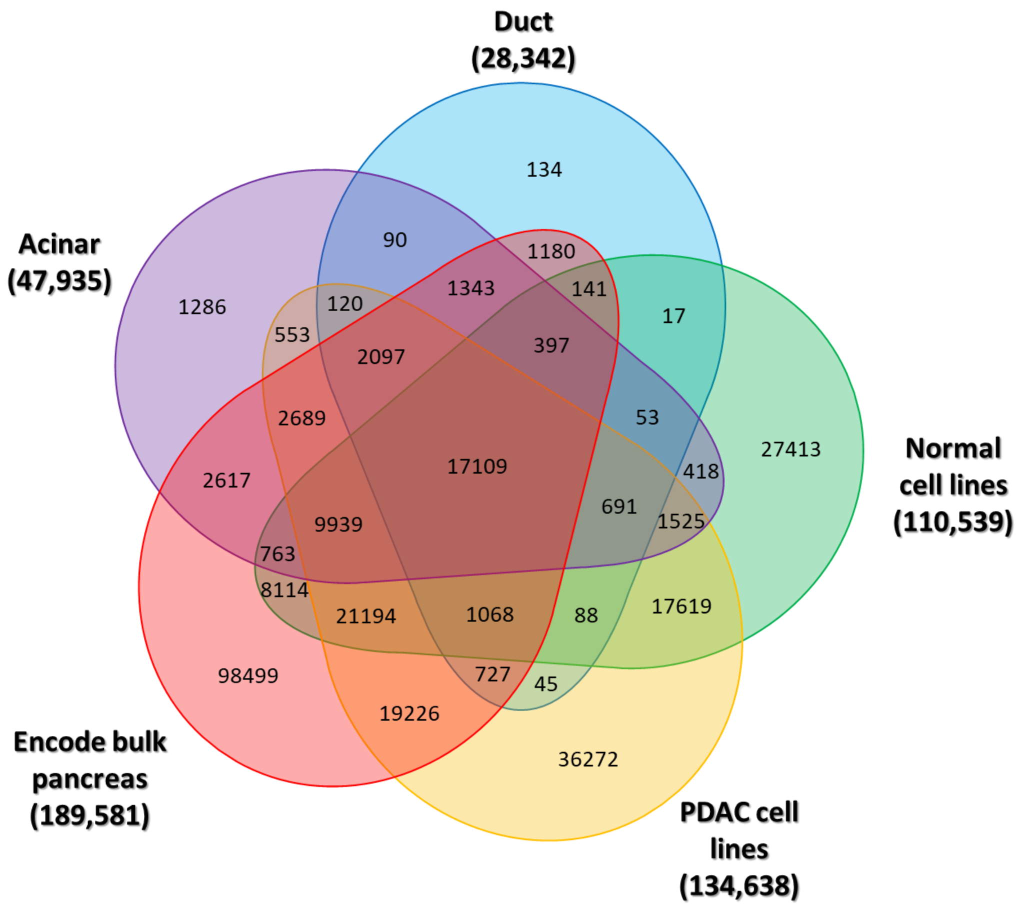


**Extended Data Fig. 5: Overlapping ACRs between this study and ENCODE bulk pancreas.** An Euler diagram depicting the tallies for overlapping ACRs between all possible combinations of sample groups as determined by the mergePeaks function in the HOMER software suite. Notably, of the 189,581 ACRs reported for these ENCODE bulk pancreas samples, 91,082 (48.0%) overlapped the ACRs used for this study while the ENCODE bulk pancreas samples missed 86,324 ACRs annotated in the pancreatic cell lines and purified acinar and duct cells.

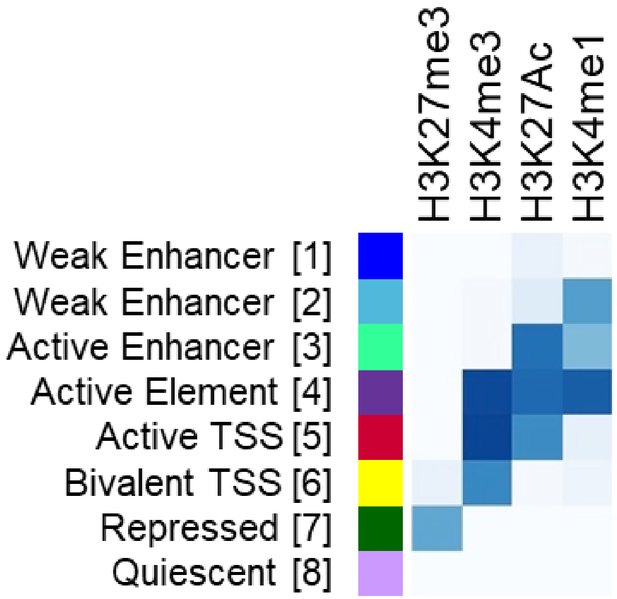


**Extended Data Fig. 6: ChromHMM states.** Heatmap summarizing the relative contributions of ChIP-seq histone modification mark (HMM) enrichments to state assignment in the ChromHMM 8-state model trained on the pancreatic cell lines and purified pancreatic acinar and duct cells as detailed in the Online Methods.

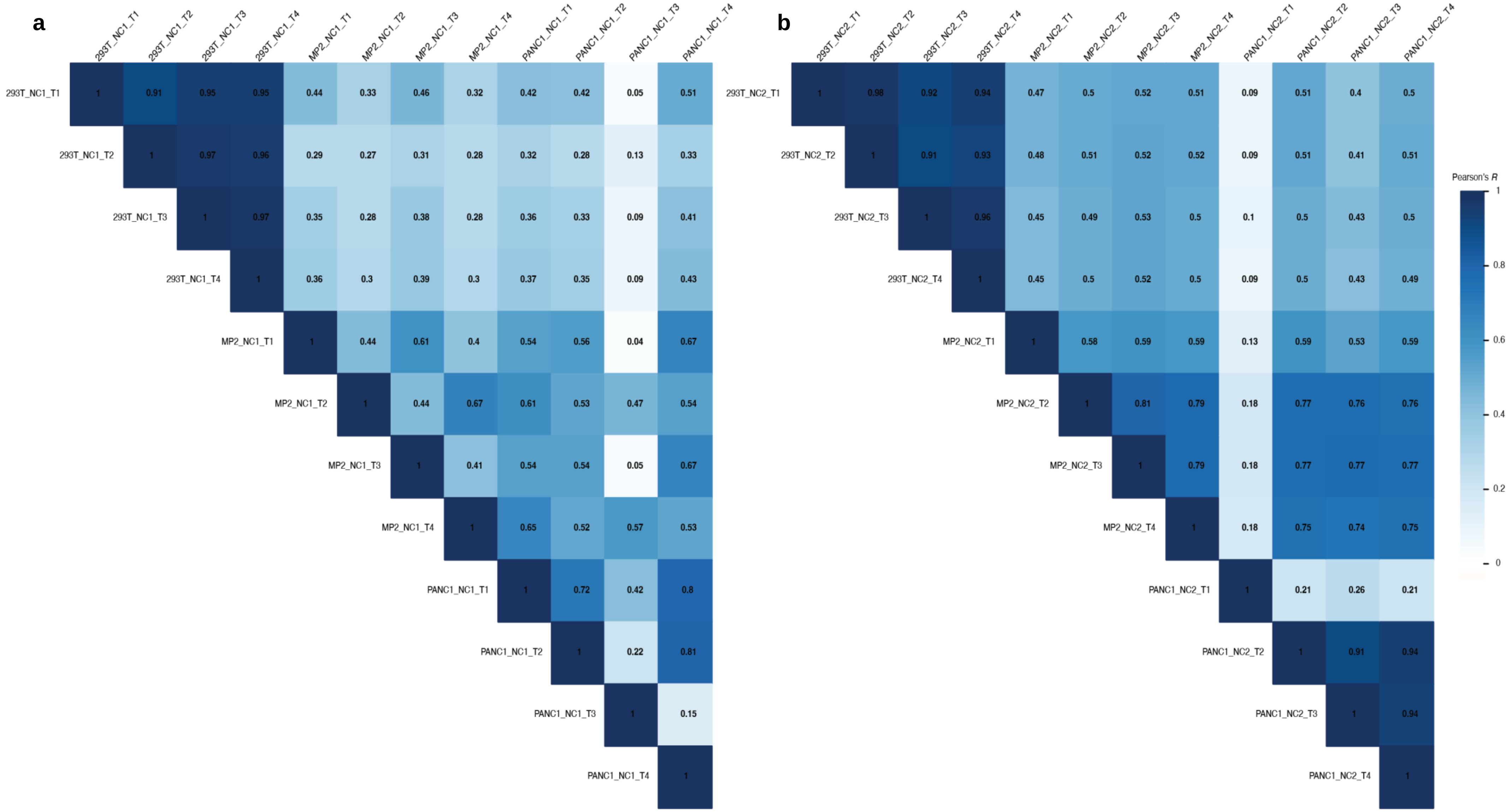


**Extended Data Fig. 7: Inter-replicate Pearson correlation of normalized RNA/DNA TPM ratios for all detected barcodes in MPRA Pool 1 (a) and Pool 2 (b) across HEK293T, PANC-1, and MIA PaCa-2 cells.** Note that one PANC-1 replicate from each MPRA pool (T3 from NC1 and T1 from NC2) were determined to be outliers by $R < 0.4$ and were removed from subsequent analysis.

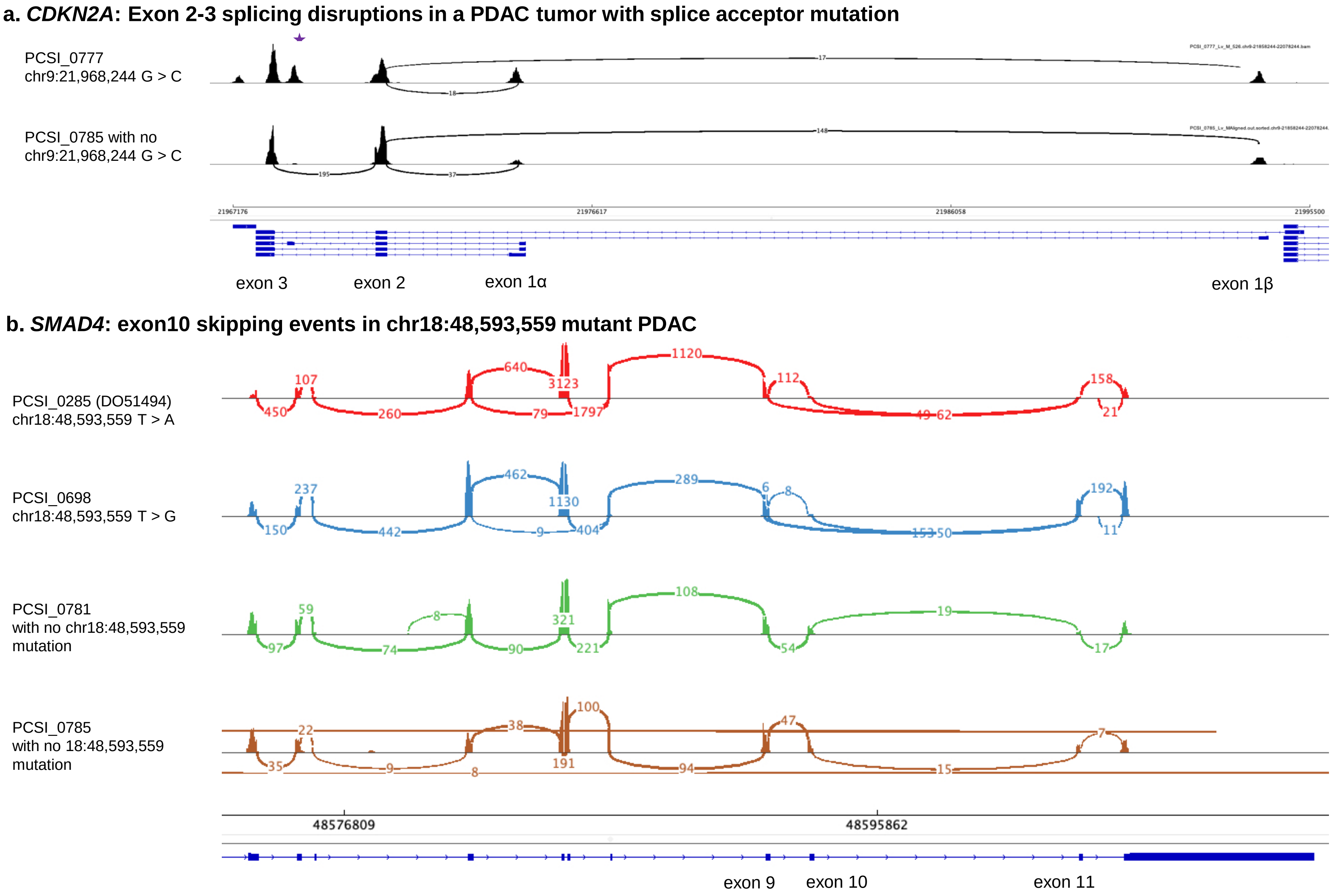


**Extended Data Fig. 8: *CDKN2A* (a) and *SMAD4* (b) splicing patterns in tumors with splicing mutations validation using RNA-seq. a,** RNA-seq data showing CDKN2A exon3-2 normal junction was disrupted in a patient (PCSI_0777) with a chr9:21,968,244 (G > C) mutation while no disruption in patient (PCSI_0785) without chr9:21,968,244 (G > C) mutation. **b**, RNA-seq data showing CDKN2A exon10 skipping events in 2 patients (PCSI_0285 and PCSI_0698) with chr18:48,593,559 (T > A/G) mutations while no skipping events in 2 patients (PCSI_0781 and PCSI_0785) without chr18:48,593,559 mutation.

c

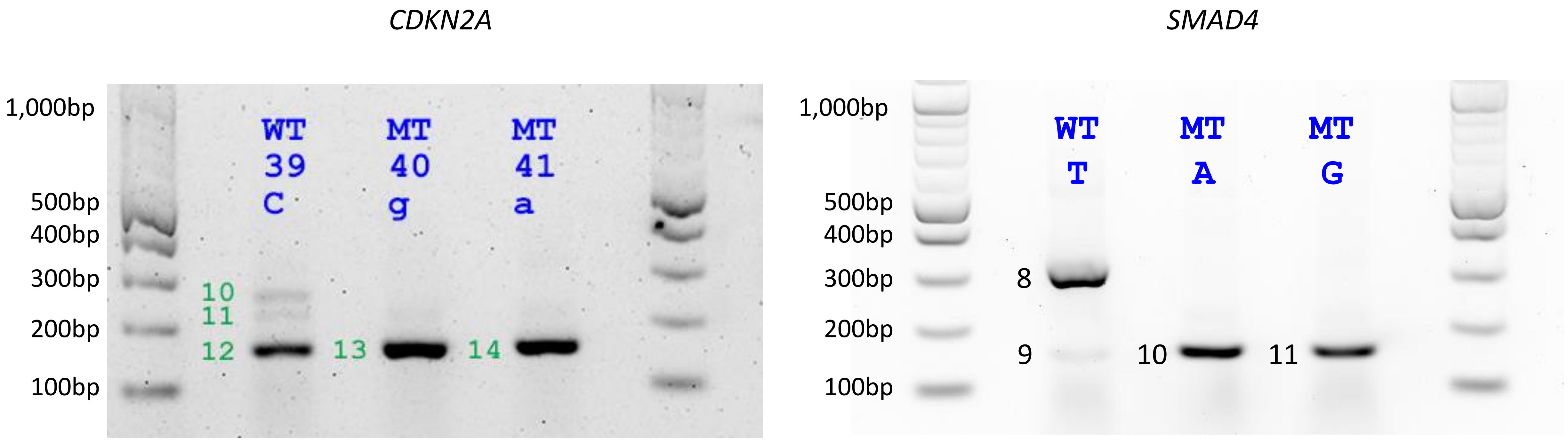


**Extended Data Fig. 8c. Functional assessment of splicing mutations in *CDKN2A* and *SMAD4* in PANC-1 cells.** Left panel: Exon-trap validation for NCSMs in *CDKN2A* at chr9:21,968,244 (G > C and G > T listed at C > G and C > A) in PANC-1 cells. Right panel: Exon-trap validation for the two NCSMs in *SMAD4* at chr18:48,593,559 (T > A and T > G) in PANC-1 cells. Note absence of bands 10 and 11 for *CDKN2A* (left) and band 8 for *SMAD4* in (right) with the mutated alleles.

a

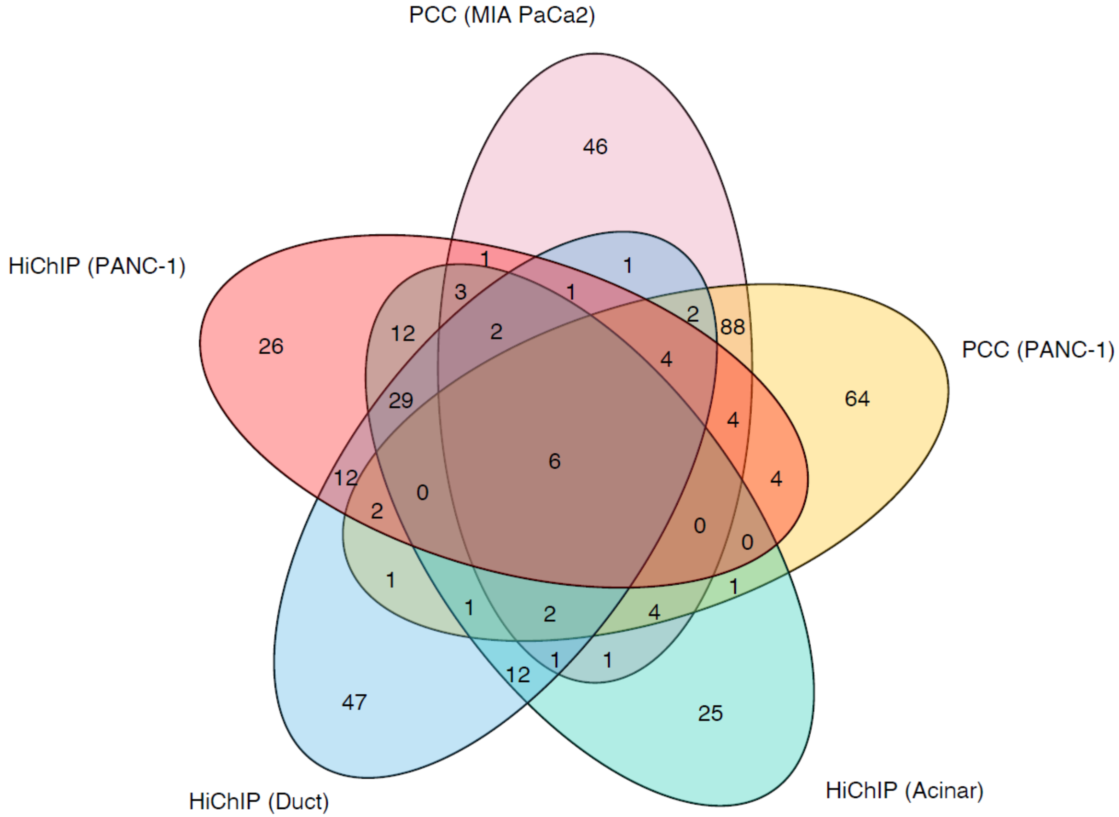


b.

| Noncoding mutation (hg38) | Target gene |
|---|---|
| chr1:149,886,685 G > C<br>chr1:149,886,706 T > C | *MTMR11*<br>*HIST2H2BC*<br>*BOLA1*<br>*ANP32E* |
| chr11:62,841,579 C > G<br>chr11:62,841,589 C > T<br>chr11:62,841,782 G > A<br>chr11:62,841,790 C > A | *SLC3A2* |
| chr9:1359,48,409 AG > -<br>chr9:135,948,424 T > G<br>chr9:133,075,338 A > G<br>chr9:133,075,366 G > C | *RALGDS* |

**Extended Data Fig. 9: Chromatin interactions between ACRs/HMMs and target genes inferred by HiChIP and PCC data (a)**. **a**. Venn diagram showing unique and shared target genes inferred by HiChIP and PCC in different cell lines. **b**. The 10 NCSMs and corresponding 6 common target genes observed in both the HiChIP and PCC data.

PANC-1 cells

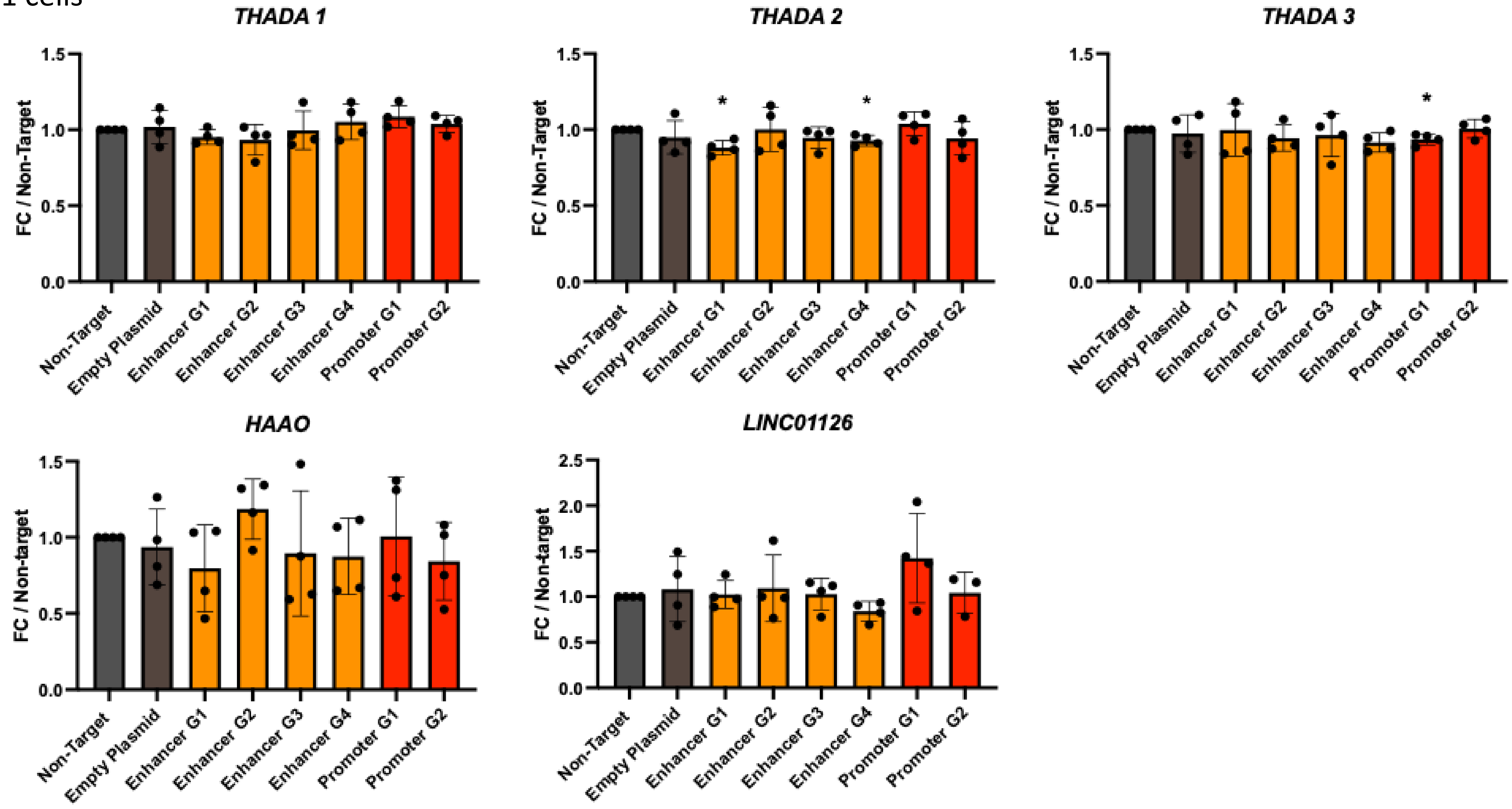


**Extended Data Fig. 10a: CRISPRi results for genes with chromatin interactions at chr2p21 near *ZFP36L2*.** Gene expression in PANC-1 cells was assessed after KD with four CRISPRi guides targeting the enhancer downstream of *ZFP36L2* (G1 – G4) as well as two *ZFP36L2* promoter guides (Promoter G1 and G2). The non-target guide is within the same topologically associating domain (TAD). Guides G1 and G4 showed significant reduction of THADA_2 expression.

MIA PaCa-2

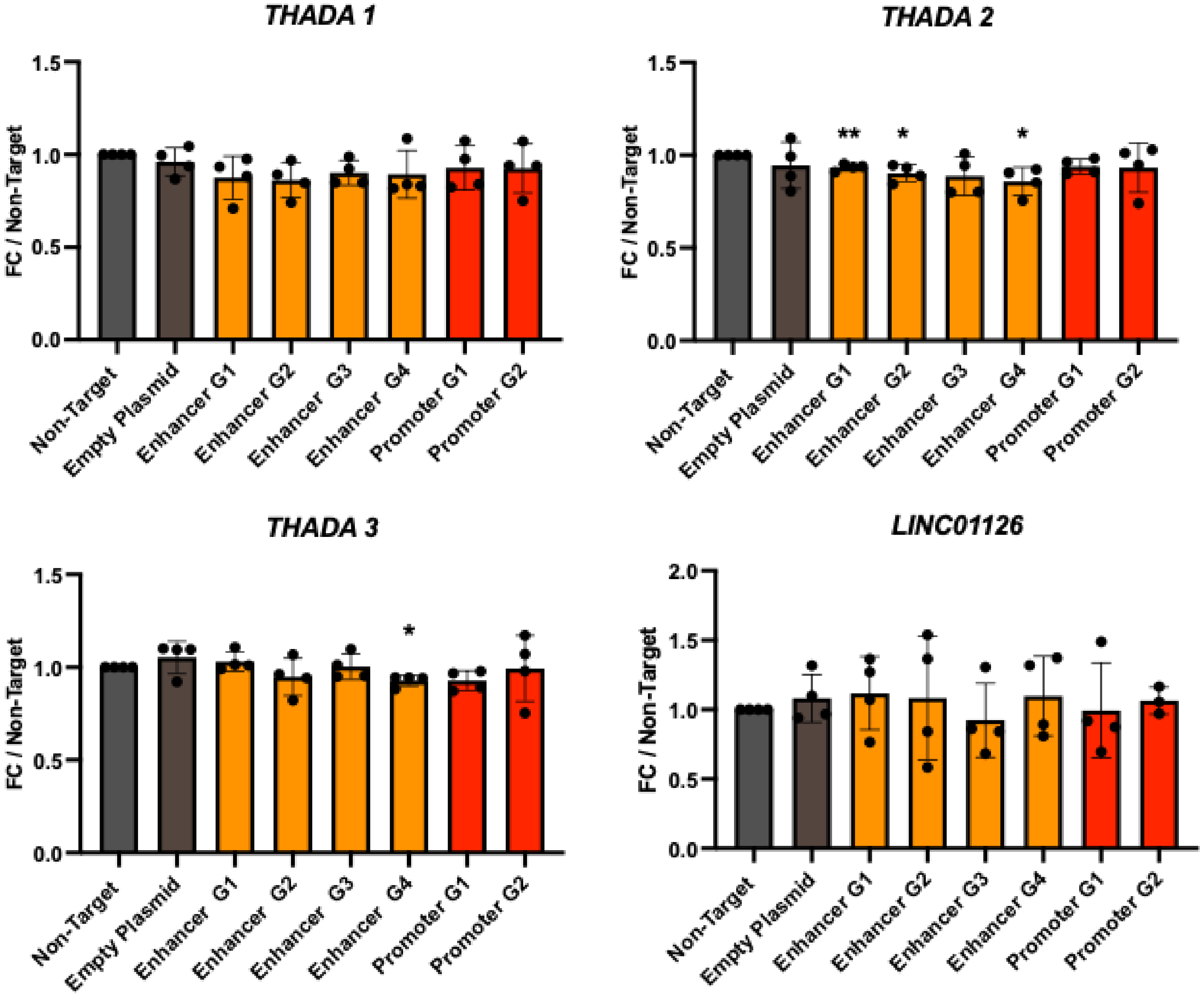


**Extended Data Fig. 10b: CRISPRi results for genes with chromatin interactions at chr2p21 near *ZFP36L2*.** Gene expression in MIA PaCa-2 cells was assessed after KD with four CRISPRi guides targeting the enhancer downstream of *ZFP36L2* (G1 – G4) as well as two *ZFP36L2* promoter guides (Promoter G1 and G2). The non-target guide is within the same topologically associating domain (TAD). Guides G1, G2 and G4 showed significant reduction of THADA_2 expression. HAAO was not expressed in this cell line.

a

Numbers of NCSMs in the 7 noncoding regions

| intergenic regions | introns | ncRNA introns | 3' UTRs | ncRNA exons | 5' UTRs | splicing mutations |
|---|---|---|---|---|---|---|
| 2,063,280 | 1,085,173 | 226,365 | 26,537 | 11,841 | 5,375 | 603 |

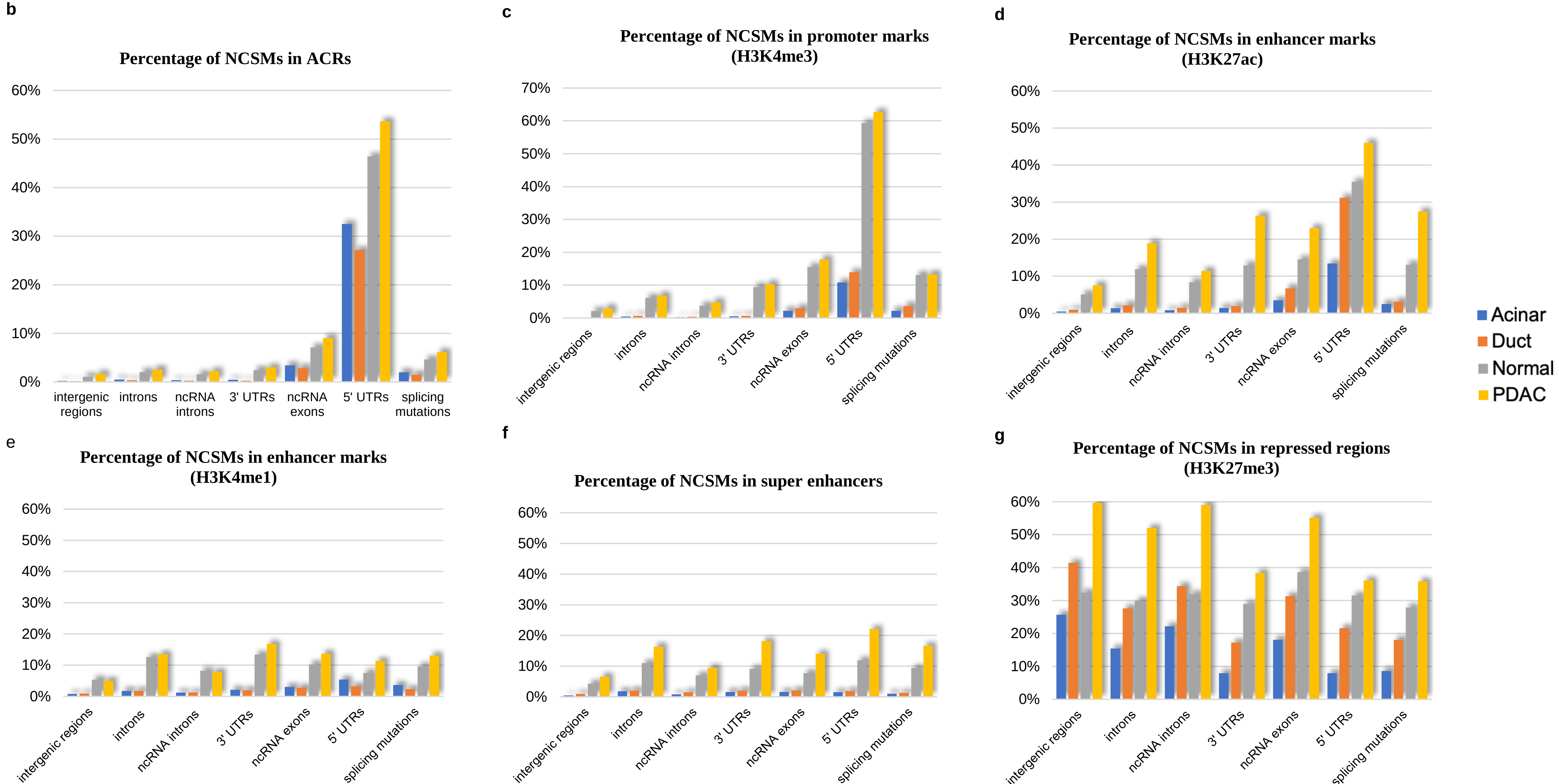


**Extended Data Fig. 11: Percentages of NCSMs in ACRs/HMMs from acinar, ductal, normal- and tumor-derived cell lines.** a, Numbers of NCSMs in the 7 noncoding regions: intergenic regions, introns, ncRNA introns, 3′UTRs, 5′UTRs, ncRNA exons and splicing mutations. **b**, Percentage of NCSMs in ACRs. **c**, Percentage of NCSMs in H3K4me3 marks. **d**, Percentage of NCSMs in H3K27ac marks. **e**, Percentage of NCSMs in marks (H3K4me1). **f**, Percentage of NCSMs in super-enhancers**. g**, Percentage of NCSMs in repressed regions (H3K27me3).

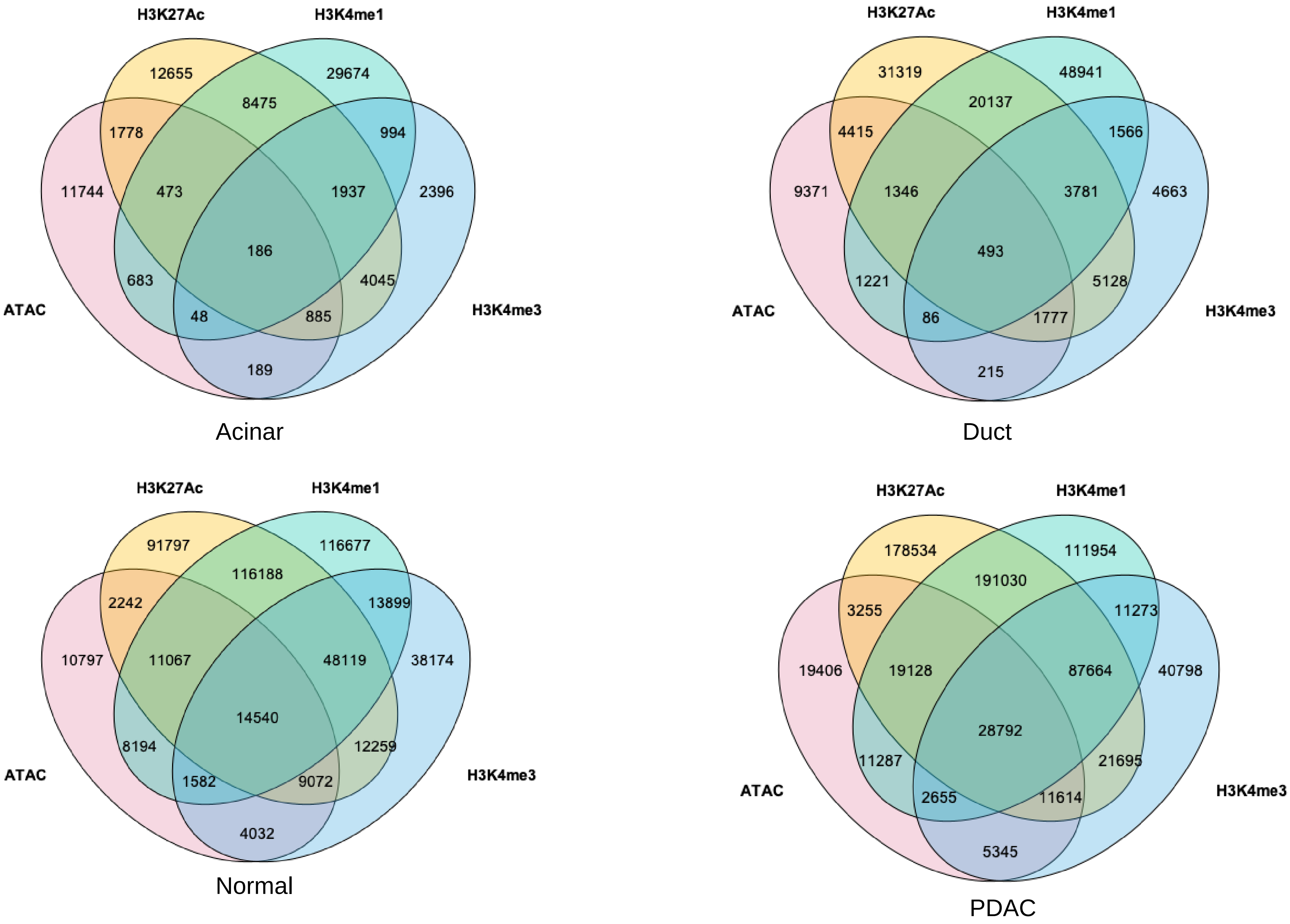


**Extended Data Fig. 12: NCSMs located in ACRs/HMMs from purified pancreatic acinar, ductal cells, normal-derived pancreatic cell lines, and PDAC cell lines.** Venn diagrams showing unique and shared NCSMs in ACRs/HMMs in the four types of cells.

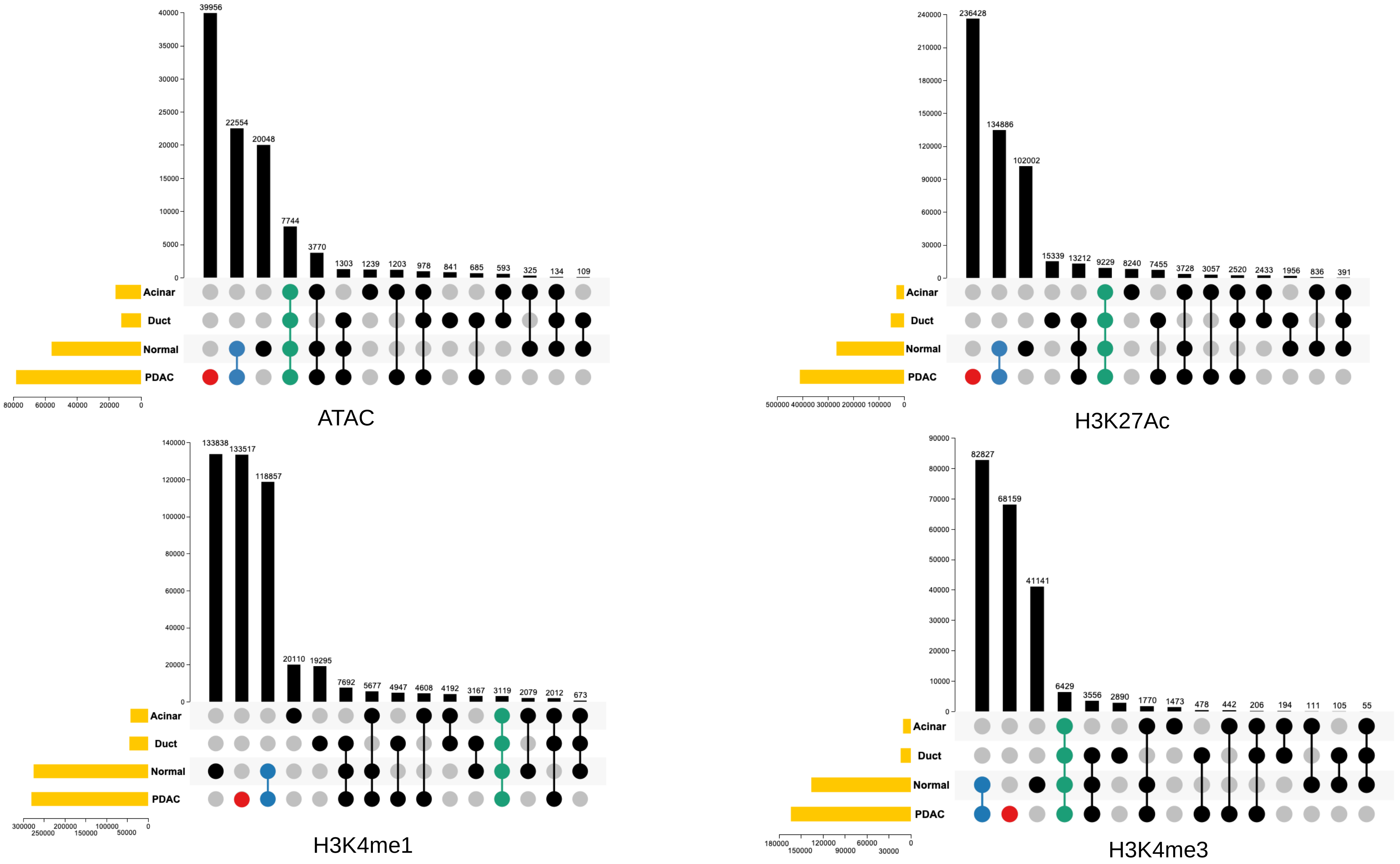


**Extended Data Fig. 13: Intersection of NCSMs in ACRs/ HMMs in purified pancreatic acinar, ductal cells, normal- and tumor-derived cell lines.** UpSet plots showing unique and shared NCSMs in the four types of cells in ACRs/HMMs.

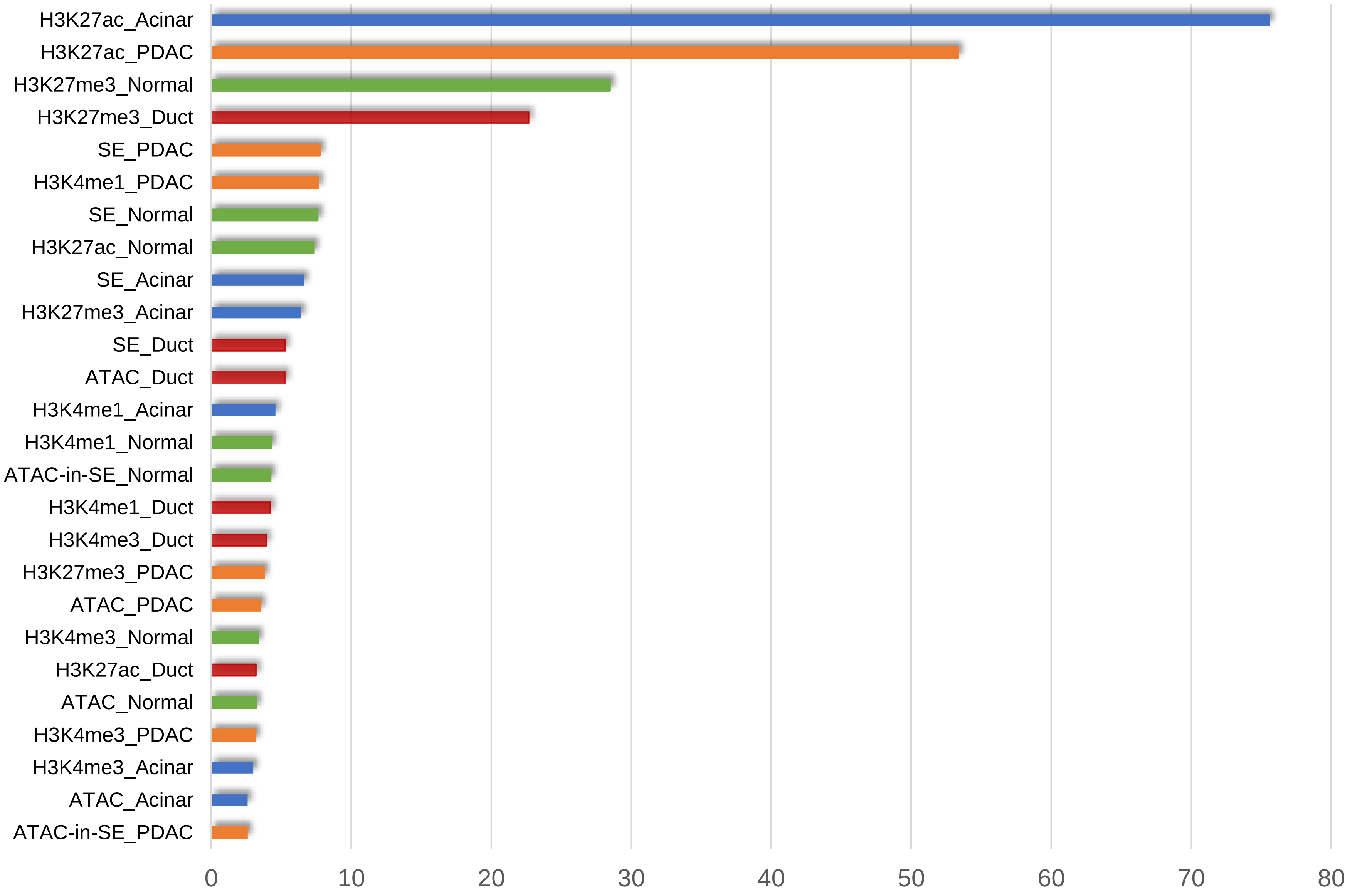


**Extended Data Fig. 14: Feature importance for background mutation rate prediction by DriverPower.** H3K27ac marks from acinar cells were the most important group of predictors for background mutation rate (BMR) using DriverPower. BMR was calculated by non-overlapped 1 megabase pair (Mbp) autosomal elements as well as training ACRs/HMMs by sampling genomic coordinates randomly. The number of mutations per element was then predicted with fivefold cross validation.
